# Development of a Real-Time Risk Model (RTRM) for Predicting In-Hospital COVID-19 Mortality

**DOI:** 10.1101/2021.04.26.21256138

**Authors:** Daniel Schlauch, Arielle M. Fisher, Jessica Correia, Xiaotong Fu, Casey Martin, Angela Junglen, Howard A. Burris, Lindsay E. Sears, Gregg Fromell, Mick Correll, Charles F. LeMaistre, Shanna A. Arnold Egloff

**Author notes:** Corresponding Author: Shanna A. Arnold Egloff, PhD, MSCI, Sarah Cannon, 1100 Dr. Martin L. King Jr. Blvd., Suite 800, Nashville, TN 37203.

## Abstract

**Background:** With over 83 million cases and 1.8 million deaths reported worldwide by the end of 2020 for SARS-CoV-2 (COVID-19), there is an urgent need to enhance identification of high-risk populations to properly evaluate therapy effectiveness with real-world evidence and improve outcomes.

**Methods:** Baseline and daily indicators were evaluated using electronic health records for 46,971 patients hospitalized with COVID-19 from 176 HCA Healthcare-affiliated hospitals, presenting from March to September 2020, to develop a real-time risk model (RTRM) of all-cause, hospitalized mortality. Patient facility, dates-of-care, clinico-demographics, comorbidities, vitals, laboratory markers, and respiratory support findings were aggregated in a logistic regression model.

**Findings:** The RTRM predicted overall mortality as well as mortality 1, 3, and 7 days in advance with an area under the receiver operating characteristic curve (AUCROC) of 0.905, 0.911, 0.905, and 0.901 respectively, significantly outperforming a combined model of age and daily modified WHO progression scale (all *p*<0.0001; AUCROC, 0.846, 0.848, 0.850, and 0.852). The RTRM delineated risk at presentation from ongoing risk associated with medical care and showed that mortality rates decreased over time due to both decreased severity and changes in care.

**Interpretation:** To our knowledge, this study is the largest of its kind to comprehensively evaluate predictors and incorporate daily risk measures of COVID-19 mortality. The RTRM validates current literature trends in mortality across time and allows direct translation for research and clinical applications.

**Research in context:** *Evidence before this study:* Due to the rapidly evolving nature of the COVID-19 pandemic, the body of evidence and published literature was considered prior to study initiation and throughout the course of the study. Although at study initiation there was a growing consensus that age and disease severity at presentation were the greatest contributors to predicting in-hospital mortality, there was less of a consensus on the key demographics, comorbidities, vitals and laboratory values. In addition, early on, most potential predictors of in-hospital mortality had been assessed by univariable analysis. In April of 2020, a systematic review of prediction studies for COVID-19 revealed that there were only 8 publications for prognosis of hospital mortality. All were deemed to have high potential for bias due to low sample size, model overfitting, vague reporting and/or insufficient follow-up. Over the duration of the study, in-hospital prediction models were published ranging from simplified scores to machine learning. There were at least 8 prediction studies that were published during the course of our own that had comparable sample size or extensive multivariable analysis with the greatest accuracy of prediction reported as 74%. Moreover, a report in December of 2020 independently validated 4 simple prediction models, with none achieving greater than an AUCROC of 0.72%. Lastly, an eight-variable score developed by a UK consortium on a comparable sample size demonstrated an AUCROC of 0.77. To our knowledge, however, none to-date have modeled daily risk beyond baseline. We frequently assessed World Health Organization (WHO) resources as well as queried both MedRXIV and PubMed with the search terms “COVID”, “prediction”, “hospital” and “mortality” to ensure we were assessing all potential predictors of hospitalized mortality. The last search was performed on January 5, 2021 with the addition of “multi”, “daily”, “real time” or “longitudinal” terms to confirm the novelty of our study. No date restrictions or language filters were applied.

*Added value of this study:* To our knowledge, this study is the largest and most geographically diverse of its kind to comprehensively evaluate predictors of in-hospital COVID-19 mortality that are available retrospectively in electronic health records and to incorporate longitudinal, daily risk measures to create risk trajectories over the entire hospital stay. Not only does our Real-Time Risk Model (RTRM) validate current literature, demonstrating reduced mortality over the course of the COVID-19 pandemic and identifying age and WHO severity as major drivers of mortality in regards to baseline characteristics, but it also outperforms a model of age and daily WHO score combined, achieving an AUCROC of 0.91 on the test set. Furthermore, the fact that the RTRM delineates risk at baseline from risk over the course of care allows more granular interpretation of the impact of various parameters on mortality risk, as demonstrated in the current study using both sex disparity and calendar epochs that were based on evolving treatment recommendations as proofs-of-principle.

*Implications of all the available evidence:* The goal of the RTRM was to create a flexible tool that could be used to assess intervention and treatment efficacy in real-world, evidence-based studies as well as provide real-time risk assessment to aid clinical decisions and resourcing with further development. Implications of this work are broad. The depth of the multi-facility, harmonized electronic health record (EHR) dataset coupled with the transparency we provide in the RTRM results provides a resource for others to interpret impact of markers of interest and utilize data that is relevant to their own studies. The RTRM will allow optimal matching in retrospective cohort studies and provide a more granular endpoint for evaluation of interventions beyond general effectiveness, such as optimal delivery, including dosing and timing, and identification of the population/s benefiting from an intervention or combination of interventions. In addition, beyond the scope of the current study, the RTRM and its resultant daily risk scores allow for flexibility in developing prediction models for other clinical outcomes, such as progression of pulmonary disease, need for invasive mechanical ventilation, and development of sepsis and/or multiorgan failure, all of which could provide a framework for real-time personalized care.

## Introduction

A cluster of pneumonia cases in Wuhan, China in December of 2019 has become a global pandemic, with over 83 million cases of severe acute respiratory syndrome coronavirus 2 (SARS-CoV-2) and 1.8 million deaths reported from COVID-19 worldwide by the end of 2020.^1-3^ Despite unprecedented vaccine efforts, the pandemic is expected to remain a persistent threat given the challenges of global deployment. Therefore, there is an urgent need to identify higher risk populations as well as more accurately measure ongoing disease severity and progression to identify effective therapies.

Moreover, due to practical challenges associated with operating and recruiting patients to randomized clinical trials, risk-adjusted retrospective cohort studies of real-world evidence have been essential to understanding treatment effects for COVID-19. These non-randomized studies, however, are inherently biased due to possible incomplete inclusion of prognostic indicators.

Given the global impact of the pandemic on healthcare system resources, there is critical need to identify patients at higher risk for developing lung and multiorgan dysfunction in real-time in order to intervene early and properly allocate resources. There is also a need to rapidly and systematically evaluate clinical interventions while accounting for robust risk adjustment, concurrent medications, and an evolving interventional landscape. Thus, considering the patient timeline and trajectory of risk throughout hospitalization is critical.

In this first proof-of-principle study, we leverage hospital data from HCA Healthcare, the largest private healthcare system in the U.S., to train and predict all cause, hospitalized mortality in a cohort of 46,971 patients. Our resultant real-time risk model (RTRM) is able to assess patient prognosis at presentation and throughout hospitalization. Moreover, the RTRM validates predictors for inclusion in retrospective, matched cohort studies for evaluating treatment effectiveness and produces a granular daily risk score that serves as a surrogate of disease progression and/or improvement. Importantly, we demonstrate the superiority of the RTRM at predicting hospitalized mortality in comparison to a combined model of age and the daily modified World Health Organization (WHO) clinical progression scale as the current benchmark measure of disease severity.^4-8^ Finally, we delineate baseline effects from responses across hospital care for calendar date epochs and for sex by comparing daily risk trajectories.

## Methods

### Regulatory and Patient Selection

This study was supported by HCA Healthcare and deemed exempt, non-human subjects research by the governing institutional review board. The design, analysis, and data interpretations were conducted independently by investigators. All authors testify to the accuracy and completeness of the data with the understanding that there may be issues in real-world records outside of our awareness. Electronic health records (EHRs) were compiled for patients hospitalized with laboratory-confirmed SARS-CoV-2 infection, representing COVID-19 disease, at 176 HCA Healthcare-affiliated facilities across the Unites States between March 2 and September 23 and were followed through October 7, 2020. A COVID-19 confirmed case was defined as a positive and/or presumptive positive SARS-CoV-2 result regardless of assay platform. A COVID-19-associated hospitalization was defined by the date of hospital presentation to the date of discharge or death; whereby, all consecutive inpatient encounters such as emergency room or holding area stays, inter- or intra-HCA-affiliated hospital transfers, and/or readmissions were considered as a single hospitalization. For patients with multiple COVID-19-associated inpatient encounters separated by greater than 36 hours, only data from the first hospitalization was used. Only those patients that presented to the hospital prior to September 23, 2020 and experienced an outcome of either “discharged alive” or “death” by October 7, 2020 were included, which was 93.8% (46,971/50,059) of the overall source population (Figure 1A). Patients that were discharged or expired on the same date of hospitalization were excluded from the analysis.

**Figure 1.**
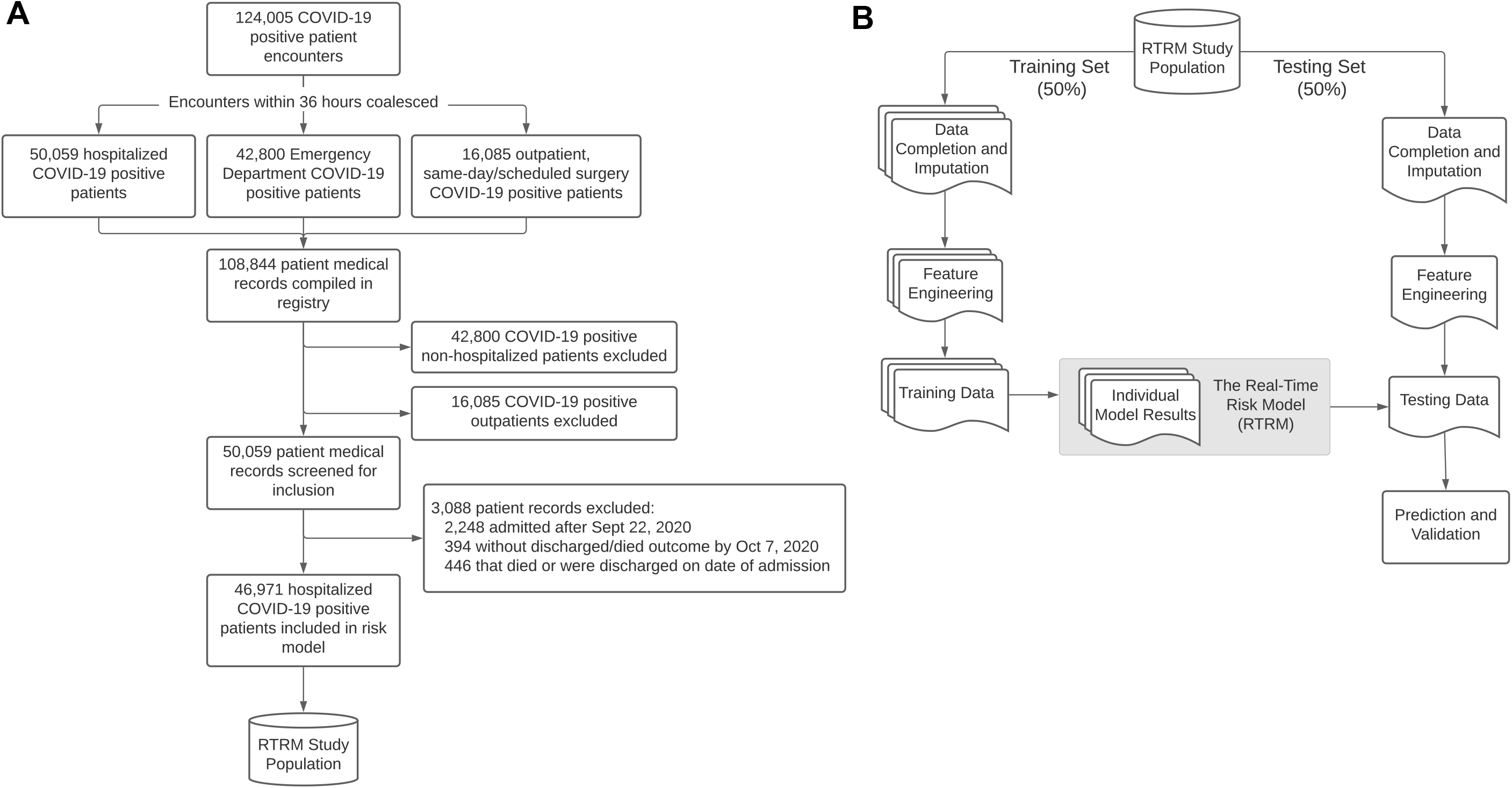
Patient Selection and Model Development Processes. Consort diagram displaying patient inclusion/exclusion and filtering criteria for defining the COVID-19 cohort of interest for evaluating hospitalized mortality (A). (B) Diagram of the data and analysis work-flow of the Real-Time Risk Model (RTRM).

### Data Collection and Definitions

Data was collected from EHRs (Epic, Cerner, Meditech) and compiled in an enterprise data warehouse. Further data processing and standardization was done using Genospace, a cloud-based biomedical data ingestion and transformation platform. This study adhered to the transparent reporting of a multivariable prediction model for individual prognosis or diagnosis (TRIPOD) reporting guidelines. Missing value handling depended on the variable and is detailed in the supplemental methods (Figure S1). Medications were purposely excluded from the RTRM to facilitate downstream assessments of treatment effectiveness.

#### Baseline Characteristics

Data collection included patient demographics, admitting and primary treating facility, smoking status, and blood type, as well as certain comorbidities documented through past and current ICD-9-CM and ICD-10-CM diagnostic codes (Table S1). Comorbidities of interest included diabetes with chronic complications, diabetes without chronic complications, hypertension, chronic ischemic heart disease, congestive heart failure, renal disease (mild or moderate), renal disease (severe), asthma or reactive airway disease, chronic pulmonary disease (excluding asthma), HIV infection, and cancer (including solid tumors and blood cancers but excluding non-melanoma skin cancer).

#### Time-Course Data

Time-course data were indexed by each day of hospitalization with baseline (t=0) defined as the date of hospital presentation (emergency room or direct admission). Longitudinal data on vitals, laboratory values, and level of respiratory support were collected throughout the course of hospitalization. Additional data processing included standard unit transformation and validation of expected values; erroneous data-points outside of plausible range were removed and considered missing. Missing data were imputed for all patient-days with no available data for a particular variable. When multiple values were present for a particular patient-day, the mean of the observed values was used.

Patients were categorized based on a modified 6-point scale, adapted from the WHO R&D Blueprint group to assess disease severity and measure clinical improvement.^4^ The modified 6-point scale, subsequently, the WHO Progression Scale (WHO PS), is as follows: 1, discharged alive; 2, hospitalized, no supplemental oxygen; 3, hospitalized, low-flow supplemental oxygen; 4, hospitalized, non-invasive or high-flow oxygen including continuous positive airway pressure (CPAP) and bi-level positive airway pressure (BIPAP); 5, hospitalized, invasive mechanical ventilation or extracorporeal membrane oxygenation (ECMO); 6, expired. Modification excluded points that could not be reliably defined by hospital EHR data. Patients were assigned a score at presentation and each day following based on their most severe status that day, except day-of-discharge, where they were assigned 1.

#### Clinical Outcomes and Complications

Clinical outcome measures were also collected, including WHO PS at discharge or death (primary outcome measure); maximum prior WHO PS; time on ventilator and time to first ventilation; intensive care unit (ICU) status, defined as receiving care in the ICU at any point during hospitalization; length of stay, defined as the time between date of presentation and death or discharge; and length of ICU stay, defined as the time between ICU admission and death or discharge from the ICU. Complications included development of acute respiratory distress syndrome (ARDS), defined as PaO2/FiO2 ratio ≤300 mmHg and a daily WHO PS of 3 or higher, as well as pneumonia, sepsis, and bacteremia based on ICD-10-CM codes (Table S2).

#### Defining Calendar Epochs

We evaluated the COVID-19 cohort as six independent date-of-hospital-presentation epochs to account for changes in treatment recommendations by the HCA Healthcare Clinical Operations Group over time in 2020. These epochs are detailed in Table 1.

**Table 1.** Definitions of six calendar epochs used to account for changes in treatment recommendations over time in 2020.

| Epoch Number | Calendar Definitions | Treatment Recommendations Summary |
| --- | --- | --- |
| Epoch 1 | March 2 – April 2, 2020 | • Consider only hydroxychloroquine without azithromycin<br>• Consider remdesivir<br>• Avoid steroids |
| Epoch 2 | April 3 – April 29, 2020 | • Consider hydroxychloroquine with azithromycin<br>• Consider tocilizumab<br>• Consider convalescent plasma |
| Epoch 3 | April 30 – May 19, 2020 | • Consider only hydroxychloroquine, no azithromycin<br>• Consider low-dose, short-course steroids in later pulmonary phase |
| Epoch 4 | May 20 – July 5, 2020 | • Do not consider hydroxychloroquine |
| Epoch 5 | July 6 – August 23, 2020 | • Steroids recommended for all patients requiring oxygen supplementation |
| Epoch 6 | August 24 – September 22, 2020 | • Consider convalescent plasma for patients with lesser severity illness* |

\* Shift in treatment recommendation for convalescent plasma from patient with more severe disease to patients with less severe disease based on the shift from the expanded access program (EAP) requirements to the emergency use authorization (EUA).

### Statistical Analysis

#### Feature Engineering

The relationship between labs, vitals, or other predictors with clinical outcomes is complex and nonlinear (Figures S2 and S3). Three derived features of each time-course variable were engineered to consider binary, continuous, and outlier-driven relationships, which included (1) the raw data itself, (2) a categorical representation of the data bucketed into 1, 10, 25, 50, 75, 90, and 99^th^ percentile cut-points, and (3) a normalized version fitting to Gaussian distribution. Two additional features were generated for longitudinal data to account for momentum or the change in values compared to the prior day (1-day delta) and two days prior (2-day delta), where values were set to 0 for patients in their first day after hospital presentation or first two days, respectively. Higher values indicate current increasing trend.

Statistical interactions across a number of features were evaluated. Details are provided in supplemental methods. Given that ∼400 features were investigated in the RTRM, candidate interaction testing was limited to those features that were clinically motivated or most significant as main effects to keep the model computationally tractable (Figure S4).

#### Data Imputation

We employed multiple imputation methods to impute missing data, which are detailed in supplemental methods and Figure S5.

#### Model Fitting and Performance

Patients were randomly split into training and testing sets with a 50:50 ratio (Figure 1B). Imputation and feature engineering were performed separately on each set following assignment. The RTRM can predict mortality for any *n*-days in advance; three *n*-days were reported (next-day, next-3-days, next-7-days) along with overall mortality by assigning a binary indicator to the corresponding days preceding death for each patient. Overall mortality at presentation was also modeled, which only assessed baseline risk. Each predictor consisted of the vector of all engineered features mapped to each patient-day. Model coefficients were estimated using logistic regression with group-wise elastic net regularization (Figure S6). Additional model fitting and bootstrap validation details are described in the supplemental methods.

Model performance was measured against age and daily WHO PS, alone and in combination, using the area under the receiver operating characteristic curve (AUCROC). Age and daily WHO PS were used to predict mortality n-days in advance in an unpenalized logistic regression model. The predictive scores for each of these reference models were evaluated in the testing set, along with the RTRM, and ROC curves were graphed and AUCROC computed for each of the n-day mortalities and for each day following hospital presentation.

## Results

A total of 46,971 hospitalized patients were included in the risk model (Figure 1A). The cohort median age was 63 years with an interquartile range (IQR) of 49-76 and was 51.6% male, 20.3% Black or African American, and 51.6% White or Caucasian (Table 2). Comorbidities most prevalent were hypertension (62.2%), diabetes (39.5%), and chronic ischemic heart disease (21.0%). Among the cohort, 15.6% expired, 33.2% received ICU care, and 15.1% received invasive mechanical ventilation and/or ECMO during hospitalization (Table 3). The median hospital length-of-stay was 6 days for discharged patients and 11 days for expired patients (Table 3).

**Table 2.**
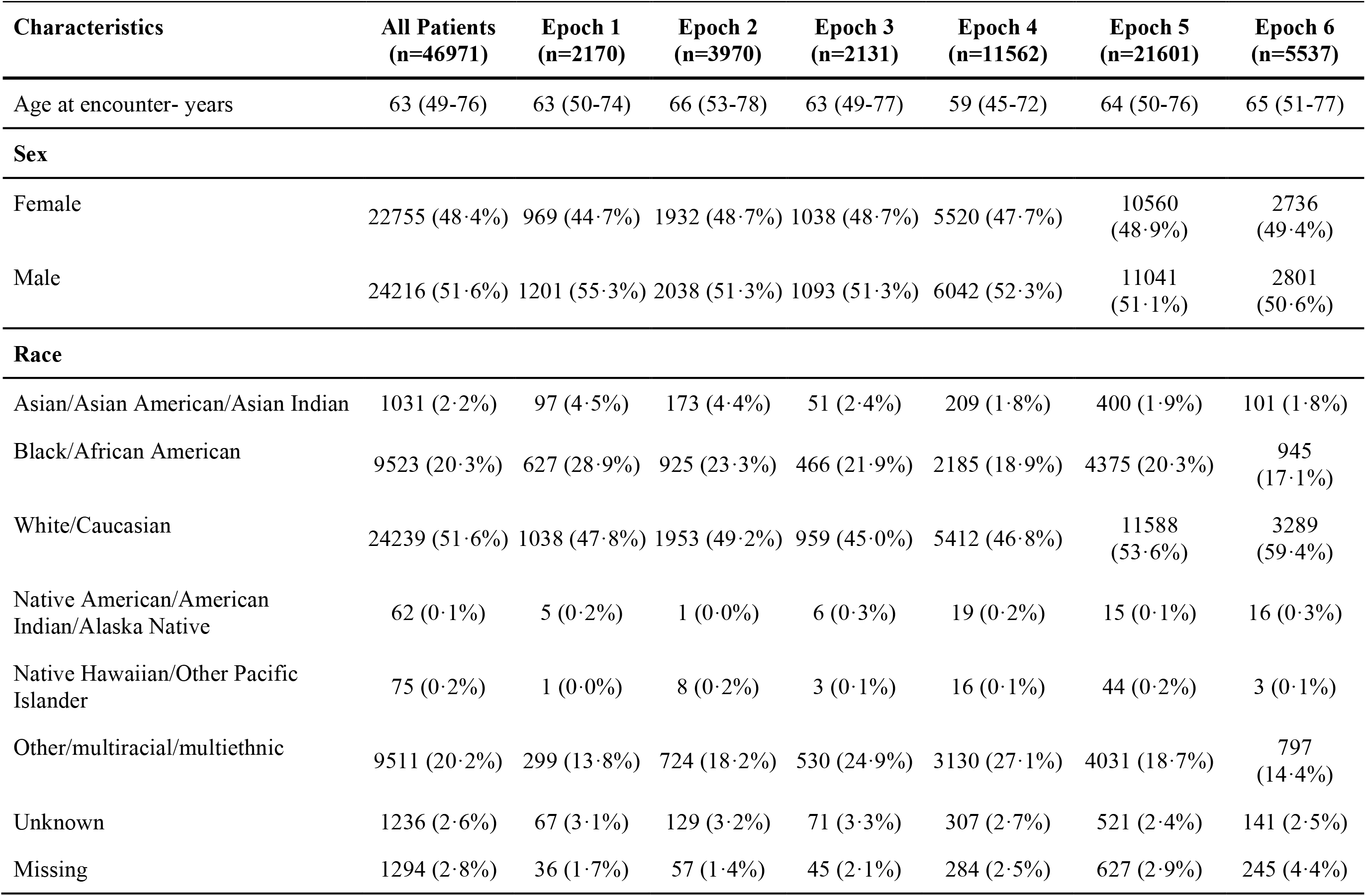
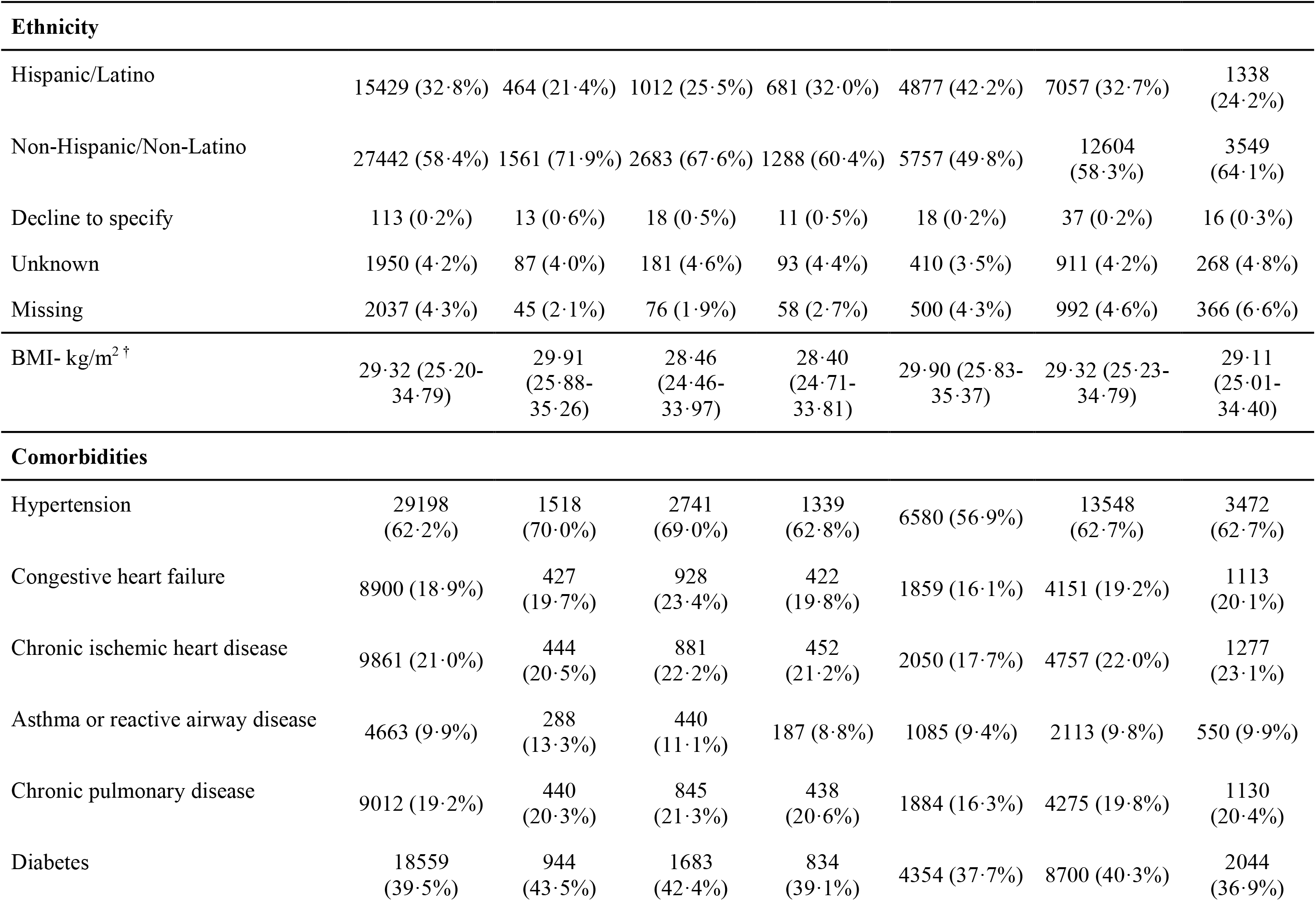
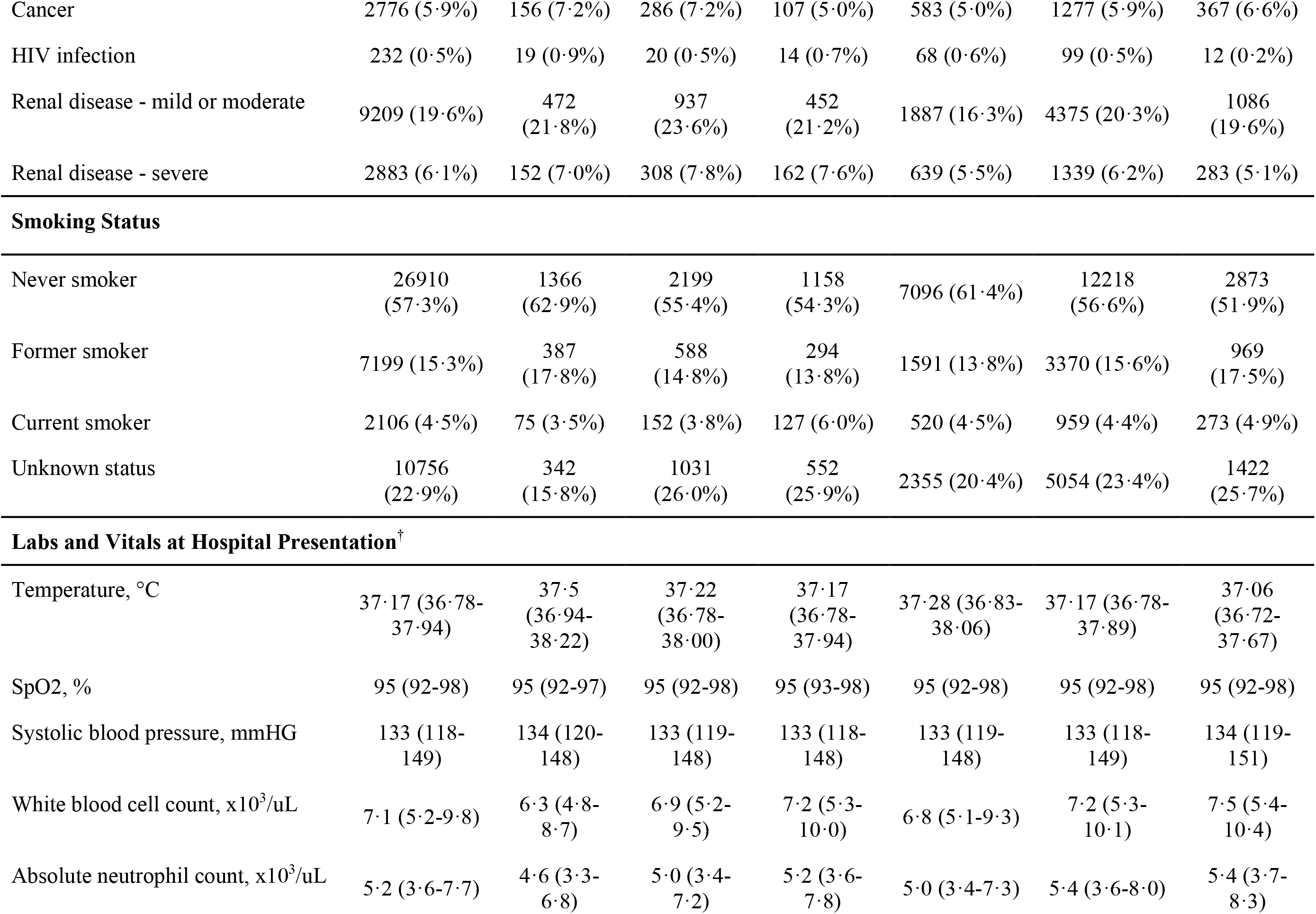
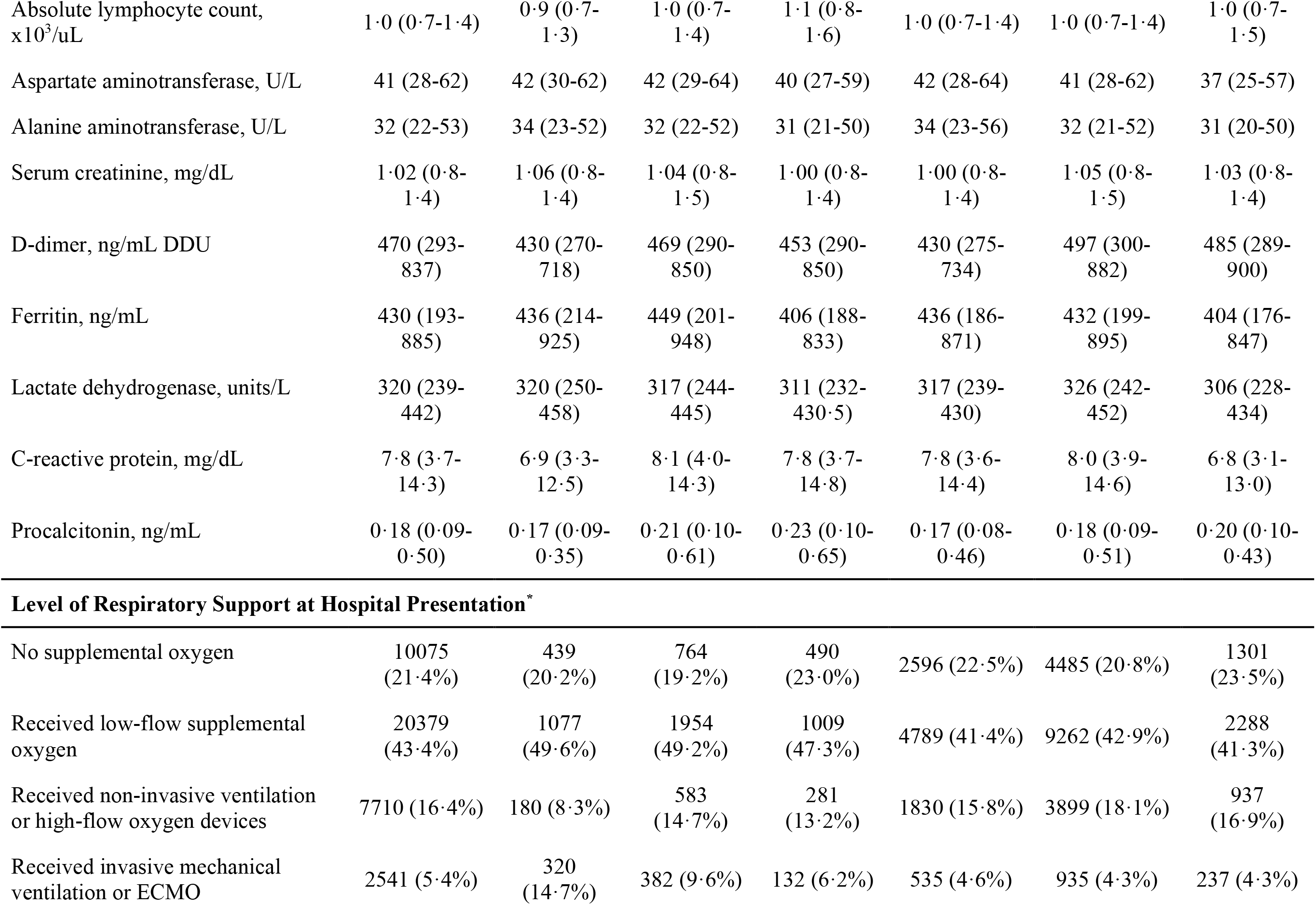
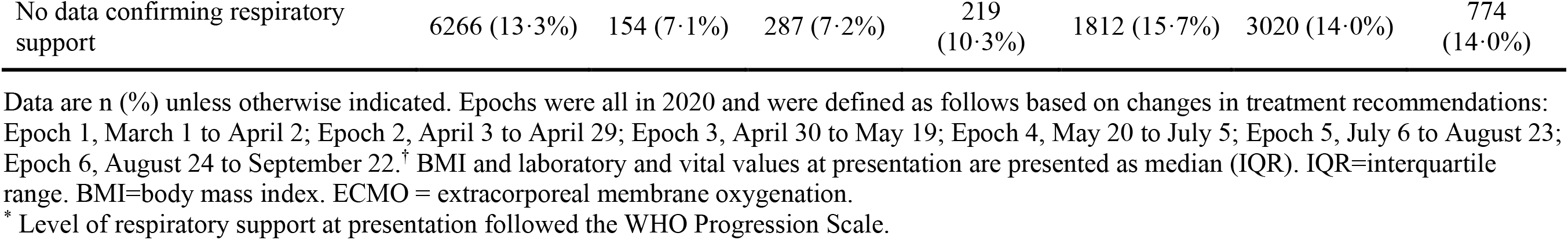
Frequencies and distributions of characteristics, comorbid conditions, initial laboratory results, and vital signs at presentation over key calendar epochs for patients hospitalized with COVID-19.

| Characteristics | All Patients<br>(n=46971) | Epoch 1<br>(n=2170) | Epoch 2<br>(n=3970) | Epoch 3<br>(n=2131) | Epoch 4<br>(n=11562) | Epoch 5<br>(n=21601) | Epoch 6<br>(n=5537) |
| --- | --- | --- | --- | --- | --- | --- | --- |
| Age at encounter- years | 63 (49-76) | 63 (50-74) | 66 (53-78) | 63 (49-77) | 59 (45-72) | 64 (50-76) | 65 (51-77) |
| **Sex** |  |  |  |  |  |  |  |
| Female | 22755 (48.4%) | 969 (44.7%) | 1932 (48.7%) | 1038 (48.7%) | 5520 (47.7%) | 10560 (48.9%) | 2736 (49.4%) |
| Male | 24216 (51.6%) | 1201 (55.3%) | 2038 (51.3%) | 1093 (51.3%) | 6042 (52.3%) | 11041 (51.1%) | 2801 (50.6%) |
| **Race** |  |  |  |  |  |  |  |
| Asian/Asian American/Asian Indian | 1031 (2.2%) | 97 (4.5%) | 173 (4.4%) | 51 (2.4%) | 209 (1.8%) | 400 (1.9%) | 101 (1.8%) |
| Black/African American | 9523 (20.3%) | 627 (28.9%) | 925 (23.3%) | 466 (21.9%) | 2185 (18.9%) | 4375 (20.3%) | 945 (17.1%) |
| White/Caucasian | 24239 (51.6%) | 1038 (47.8%) | 1953 (49.2%) | 959 (45.0%) | 5412 (46.8%) | 11588 (53.6%) | 3289 (59.4%) |
| Native American/American Indian/Alaska Native | 62 (0.1%) | 5 (0.2%) | 1 (0.0%) | 6 (0.3%) | 19 (0.2%) | 15 (0.1%) | 16 (0.3%) |
| Native Hawaiian/Other Pacific Islander | 75 (0.2%) | 1 (0.0%) | 8 (0.2%) | 3 (0.1%) | 16 (0.1%) | 44 (0.2%) | 3 (0.1%) |
| Other/multiracial/multiethnic | 9511 (20.2%) | 299 (13.8%) | 724 (18.2%) | 530 (24.9%) | 3130 (27.1%) | 4031 (18.7%) | 797 (14.4%) |
| Unknown | 1236 (2.6%) | 67 (3.1%) | 129 (3.2%) | 71 (3.3%) | 307 (2.7%) | 521 (2.4%) | 141 (2.5%) |
| Missing | 1294 (2.8%) | 36 (1.7%) | 57 (1.4%) | 45 (2.1%) | 284 (2.5%) | 627 (2.9%) | 245 (4.4%) |

| **Ethnicity** |  |  |  |  |  |  |  |
| --- | --- | --- | --- | --- | --- | --- | --- |
| Hispanic/Latino | 15429 (32·8%) | 464 (21·4%) | 1012 (25·5%) | 681 (32·0%) | 4877 (42·2%) | 7057 (32·7%) | 1338 (24·2%) |
| Non-Hispanic/Non-Latino | 27442 (58·4%) | 1561 (71·9%) | 2683 (67·6%) | 1288 (60·4%) | 5757 (49·8%) | 12604 (58·3%) | 3549 (64·1%) |
| Decline to specify | 113 (0·2%) | 13 (0·6%) | 18 (0·5%) | 11 (0·5%) | 18 (0·2%) | 37 (0·2%) | 16 (0·3%) |
| Unknown | 1950 (4·2%) | 87 (4·0%) | 181 (4·6%) | 93 (4·4%) | 410 (3·5%) | 911 (4·2%) | 268 (4·8%) |
| Missing | 2037 (4·3%) | 45 (2·1%) | 76 (1·9%) | 58 (2·7%) | 500 (4·3%) | 992 (4·6%) | 366 (6·6%) |
| BMI- kg/m<sup>2</sup> † | 29·32 (25·20-34·79) | 29·91 (25·88-35·26) | 28·46 (24·46-33·97) | 28·40 (24·71-33·81) | 29·90 (25·83-35·37) | 29·32 (25·23-34·79) | 29·11 (25·01-34·40) |
| **Comorbidities** |  |  |  |  |  |  |  |
| Hypertension | 29198 (62·2%) | 1518 (70·0%) | 2741 (69·0%) | 1339 (62·8%) | 6580 (56·9%) | 13548 (62·7%) | 3472 (62·7%) |
| Congestive heart failure | 8900 (18·9%) | 427 (19·7%) | 928 (23·4%) | 422 (19·8%) | 1859 (16·1%) | 4151 (19·2%) | 1113 (20·1%) |
| Chronic ischemic heart disease | 9861 (21·0%) | 444 (20·5%) | 881 (22·2%) | 452 (21·2%) | 2050 (17·7%) | 4757 (22·0%) | 1277 (23·1%) |
| Asthma or reactive airway disease | 4663 (9·9%) | 288 (13·3%) | 440 (11·1%) | 187 (8·8%) | 1085 (9·4%) | 2113 (9·8%) | 550 (9·9%) |
| Chronic pulmonary disease | 9012 (19·2%) | 440 (20·3%) | 845 (21·3%) | 438 (20·6%) | 1884 (16·3%) | 4275 (19·8%) | 1130 (20·4%) |
| Diabetes | 18559 (39·5%) | 944 (43·5%) | 1683 (42·4%) | 834 (39·1%) | 4354 (37·7%) | 8700 (40·3%) | 2044 (36·9%) |
| Cancer | 2776 (5.9%) | 156 (7.2%) | 286 (7.2%) | 107 (5.0%) | 583 (5.0%) | 1277 (5.9%) | 367 (6.6%) |
| HIV infection | 232 (0.5%) | 19 (0.9%) | 20 (0.5%) | 14 (0.7%) | 68 (0.6%) | 99 (0.5%) | 12 (0.2%) |
| Renal disease - mild or moderate | 9209 (19.6%) | 472 (21.8%) | 937 (23.6%) | 452 (21.2%) | 1887 (16.3%) | 4375 (20.3%) | 1086 (19.6%) |
| Renal disease - severe | 2883 (6.1%) | 152 (7.0%) | 308 (7.8%) | 162 (7.6%) | 639 (5.5%) | 1339 (6.2%) | 283 (5.1%) |
| Smoking Status |  |  |  |  |  |  |  |
| Never smoker | 26910 (57.3%) | 1366 (62.9%) | 2199 (55.4%) | 1158 (54.3%) | 7096 (61.4%) | 12218 (56.6%) | 2873 (51.9%) |
| Former smoker | 7199 (15.3%) | 387 (17.8%) | 588 (14.8%) | 294 (13.8%) | 1591 (13.8%) | 3370 (15.6%) | 969 (17.5%) |
| Current smoker | 2106 (4.5%) | 75 (3.5%) | 152 (3.8%) | 127 (6.0%) | 520 (4.5%) | 959 (4.4%) | 273 (4.9%) |
| Unknown status | 10756 (22.9%) | 342 (15.8%) | 1031 (26.0%) | 552 (25.9%) | 2355 (20.4%) | 5054 (23.4%) | 1422 (25.7%) |
| Labs and Vitals at Hospital Presentation<sup>†</sup> |  |  |  |  |  |  |  |
| Temperature, °C | 37.17 (36.78-37.94) | 37.5 (36.94-38.22) | 37.22 (36.78-38.00) | 37.17 (36.78-37.94) | 37.28 (36.83-38.06) | 37.17 (36.78-37.89) | 37.06 (36.72-37.67) |
| SpO2, % | 95 (92-98) | 95 (92-97) | 95 (92-98) | 95 (93-98) | 95 (92-98) | 95 (92-98) | 95 (92-98) |
| Systolic blood pressure, mmHG | 133 (118-149) | 134 (120-148) | 133 (119-148) | 133 (118-148) | 133 (119-148) | 133 (118-149) | 134 (119-151) |
| White blood cell count, x10<sup>3</sup>/uL | 7.1 (5.2-9.8) | 6.3 (4.8-8.7) | 6.9 (5.2-9.5) | 7.2 (5.3-10.0) | 6.8 (5.1-9.3) | 7.2 (5.3-10.1) | 7.5 (5.4-10.4) |
| Absolute neutrophil count, x10<sup>3</sup>/uL | 5.2 (3.6-7.7) | 4.6 (3.3-6.8) | 5.0 (3.4-7.2) | 5.2 (3.6-7.8) | 5.0 (3.4-7.3) | 5.4 (3.6-8.0) | 5.4 (3.7-8.3) |
| Absolute lymphocyte count, x10<sup>3</sup>/uL | 1·0 (0·7-1·4) | 0·9 (0·7-1·3) | 1·0 (0·7-1·4) | 1·1 (0·8-1·6) | 1·0 (0·7-1·4) | 1·0 (0·7-1·4) | 1·0 (0·7-1·5) |
| Aspartate aminotransferase, U/L | 41 (28-62) | 42 (30-62) | 42 (29-64) | 40 (27-59) | 42 (28-64) | 41 (28-62) | 37 (25-57) |
| Alanine aminotransferase, U/L | 32 (22-53) | 34 (23-52) | 32 (22-52) | 31 (21-50) | 34 (23-56) | 32 (21-52) | 31 (20-50) |
| Serum creatinine, mg/dL | 1·02 (0·8-1·4) | 1·06 (0·8-1·4) | 1·04 (0·8-1·5) | 1·00 (0·8-1·4) | 1·00 (0·8-1·4) | 1·05 (0·8-1·5) | 1·03 (0·8-1·4) |
| D-dimer, ng/mL DDU | 470 (293-837) | 430 (270-718) | 469 (290-850) | 453 (290-850) | 430 (275-734) | 497 (300-882) | 485 (289-900) |
| Ferritin, ng/mL | 430 (193-885) | 436 (214-925) | 449 (201-948) | 406 (188-833) | 436 (186-871) | 432 (199-895) | 404 (176-847) |
| Lactate dehydrogenase, units/L | 320 (239-442) | 320 (250-458) | 317 (244-445) | 311 (232-430·5) | 317 (239-430) | 326 (242-452) | 306 (228-434) |
| C-reactive protein, mg/dL | 7·8 (3·7-14·3) | 6·9 (3·3-12·5) | 8·1 (4·0-14·3) | 7·8 (3·7-14·8) | 7·8 (3·6-14·4) | 8·0 (3·9-14·6) | 6·8 (3·1-13·0) |
| Procalcitonin, ng/mL | 0·18 (0·09-0·50) | 0·17 (0·09-0·35) | 0·21 (0·10-0·61) | 0·23 (0·10-0·65) | 0·17 (0·08-0·46) | 0·18 (0·09-0·51) | 0·20 (0·10-0·43) |
| Level of Respiratory Support at Hospital Presentation\* |  |  |  |  |  |  |  |
| No supplemental oxygen | 10075<br>(21·4%) | 439<br>(20·2%) | 764<br>(19·2%) | 490<br>(23·0%) | 2596 (22·5%) | 4485 (20·8%) | 1301<br>(23·5%) |
| Received low-flow supplemental oxygen | 20379<br>(43·4%) | 1077<br>(49·6%) | 1954<br>(49·2%) | 1009<br>(47·3%) | 4789 (41·4%) | 9262 (42·9%) | 2288<br>(41·3%) |
| Received non-invasive ventilation or high-flow oxygen devices | 7710 (16·4%) | 180 (8·3%) | 583<br>(14·7%) | 281<br>(13·2%) | 1830 (15·8%) | 3899 (18·1%) | 937<br>(16·9%) |
| Received invasive mechanical ventilation or ECMO | 2541 (5·4%) | 320<br>(14·7%) | 382 (9·6%) | 132 (6·2%) | 535 (4·6%) | 935 (4·3%) | 237 (4·3%) |
| No data confirming respiratory support | 6266 (13·3%) | 154 (7·1%) | 287 (7·2%) | 219 (10·3%) | 1812 (15·7%) | 3020 (14·0%) | 774 (14·0%) |

Data are n (%) unless otherwise indicated. Epochs were all in 2020 and were defined as follows based on changes in treatment recommendations: Epoch 1, March 1 to April 2; Epoch 2, April 3 to April 29; Epoch 3, April 30 to May 19; Epoch 4, May 20 to July 5; Epoch 5, July 6 to August 23; Epoch 6, August 24 to September 22.<sup>†</sup> BMI and laboratory and vital values at presentation are presented as median (IQR). IQR=interquartile range. BMI=body mass index. ECMO = extracorporeal membrane oxygenation.
\* Level of respiratory support at presentation followed the WHO Progression Scale.

**Table 3.** Frequencies and distributions of severity indices- complications- and outcome measures over key calendar epochs for patients hospitalized with COVID-19.

| Clinical measure | All Patients<br>(n=46971) | Epoch 1<br>(n=2170) | Epoch 2<br>(n=3970) | Epoch 3<br>(n=2131) | Epoch 4<br>(n=11562) | Epoch 5<br>(n=21601) | Epoch 6<br>(n=5537) |
| --- | --- | --- | --- | --- | --- | --- | --- |
| Expired | 7327 (15.6%) | 415 (19.1%) | 844 (21.3%) | 330 (15.5%) | 1637 (14.2%) | 3418<br>(15.8%) | 683<br>(12.3%) |
| Received invasive mechanical ventilation | 7095 (15.1%) | 665 (30.6%) | 840 (21.2%) | 328 (15.4%) | 1690 (14.6%) | 2984<br>(13.8%) | 588<br>(10.6%) |
| Time to invasive mechanical ventilation, days<sup>§</sup> | 3 (0-8) | 1 (0-3) | 1 (0-4) | 2 (0-7) | 4 (0-10) | 4 (0-10) | 2 (0-8) |
| Received ICU care | 15602<br>(33.2%) | 1006<br>(46.4%) | 1691<br>(42.6%) | 912<br>(42.8%) | 4110<br>(35.5%) | 6225<br>(28.8%) | 1658<br>(29.9%) |
| Length ICU stay, days<sup>†</sup> | 5.53 (2.19-11.77) | 6.96 (3.21-14.56) | 6.74 (2.72-13.38) | 5.78 (2.44-11.88) | 5.33 (2.20-11.84) | 5.26 (2.02-11.59) | 4.73 (1.87-9.69) |
| Length of stay, days<sup>†</sup> |  |  |  |  |  |  |  |
| Discharged | 6 (3-11) | 7 (3-15) | 7 (4-15) | 7 (3-14) | 6 (3-12) | 6 (3-11) | 5 (3-8) |
| Expired | 11 (6-19) | 9 (5- 16) | 9 (4.75-17) | 11 (5.25-19) | 13 (6-22) | 12 (6-19) | 10 (4-15) |
| **Level of Respiratory Support at Most Severe\*** |  |  |  |  |  |  |  |
| No supplemental oxygen | 6790 (14.5%) | 230 (10.6%) | 443 (11.2%) | 304 (14.3%) | 1757 (15.2%) | 3073<br>(14.2%) | 983<br>(17.8%) |
| Received low-flow supplemental oxygen | 19070 (40.6%) | 888 (40.9%) | 1691 (42.6%) | 901 (42.3%) | 4726 (40.9%) | 8723<br>(40.4%) | 2141<br>(38.7%) |
| Received non-invasive ventilation or high-flow oxygen devices | 11266 (24.0%) | 324 (14.9%) | 855 (21.5%) | 480 (22.5%) | 2722 (23.5%) | 5532<br>(25.6%) | 1353<br>(24.4%) |
| Received invasive mechanical ventilation or ECMO | 7102 (15·1%) | 665 (30·6%) | 841 (21·2%) | 329 (15·4%) | 1693 (14·6%) | 2986 (13·8%) | 588 (10·6%) |
| No data confirming respiratory support | 2743 (5·8%) | 63 (2·9%) | 140 (3·5%) | 117 (5·5%) | 664 (5·7%) | 1287 (6·0%) | 472 (8·5%) |
| **Complications** |  |  |  |  |  |  |  |
| ARDS | 12648 (26·9%) | 764 (35·2%) | 1165 (29·3%) | 537 (25·2%) | 3166 (27·4%) | 5802 (26·9%) | 1214 (21·9%) |
| Bacteremia | 388 (0·8%) | 15 (0·7%) | 28 (0·7%) | 19 (0·9%) | 78 (0·7%) | 198 (0·9%) | 50 (0·9%) |
| Bacterial pneumonia | 2032 (4·3%) | 146 (6·7%) | 218 (5·5%) | 86 (4·0%) | 468 (4·0%) | 881 (4·1%) | 233 (4·2%) |
| Sepsis (excluding severe sepsis) | 10611 (22·6%) | 504 (23·2%) | 918 (23·1%) | 524 (24·6%) | 2691 (23·3%) | 4784 (22·1%) | 1190 (21·5%) |
| Severe sepsis | 8456 (18·0%) | 565 (26·0%) | 981 (24·7%) | 411 (19·3%) | 1958 (16·9%) | 3649 (16·9%) | 892 (16·1%) |
| Viral/other/unspecified pneumonia | 36388 (77·5%) | 1850 (85·3%) | 3208 (80·8%) | 1614 (75·7%) | 8817 (76·3%) | 16 905 (78·3%) | 3994 (72·1%) |

Data are n (%) unless otherwise indicated. Epochs were all in 2020 and were defined as follows based on changes in treatment recommendations: Epoch 1, March 1 to April 2; Epoch 2, April 3 to April 29; Epoch 3, April 30 to May 19; Epoch 4, May 20 to July 5; Epoch 5, July 6 to August 23; Epoch 6, August 24 to September 22. IQR=interquartile range. ECMO = extracorporeal membrane oxygenation. ARDS=acute respiratory distress syndrome.
§ Time to first episode of mechanical ventilation is presented as median (IQR).
† Length of stay and time to outcomes are presented as median (IQR).
\* Level of respiratory support at most severe followed the WHO Progression Scale.

### RTRM performance against benchmark predictors

The RTRM demonstrated strong performance in predicting mortality in real-time. For each day a test set patient remained hospitalized, the RTRM produced predictions for outcomes of next-day, next-3-days, next-7-days, and overall mortality, which resulted in an AUCROC of 0.911, 0.905, 0.901, and 0.905, respectively (Figure 2A-D). We compared the RTRM result to the performance of two benchmarks of COVID-19 mortality, age and the daily WHO PS.^4-6^ The AUCROCs were significantly lower for all models using either age alone (all *p*<0.0001; AUCROCs, 0.653, 0.656, 0.658, and 0.653) or the daily WHO PS alone (all *p*<0.0001; AUCROCs, 0.804, 0.803, 0.801, and 0.792; Table S3 and Figure 2A-D). Importantly, the RTRM significantly outperformed the model combining both age and the daily WHO PS (all *p*<0.0001; AUCROCs, 0.848, 0.850, 0.852, and 0.901; Table S3 and Figure 2A-D). This consistent outperformance across each outcome highlights the strength of the RTRM to predict mortality in the days preceding death.

**Figure 2:**
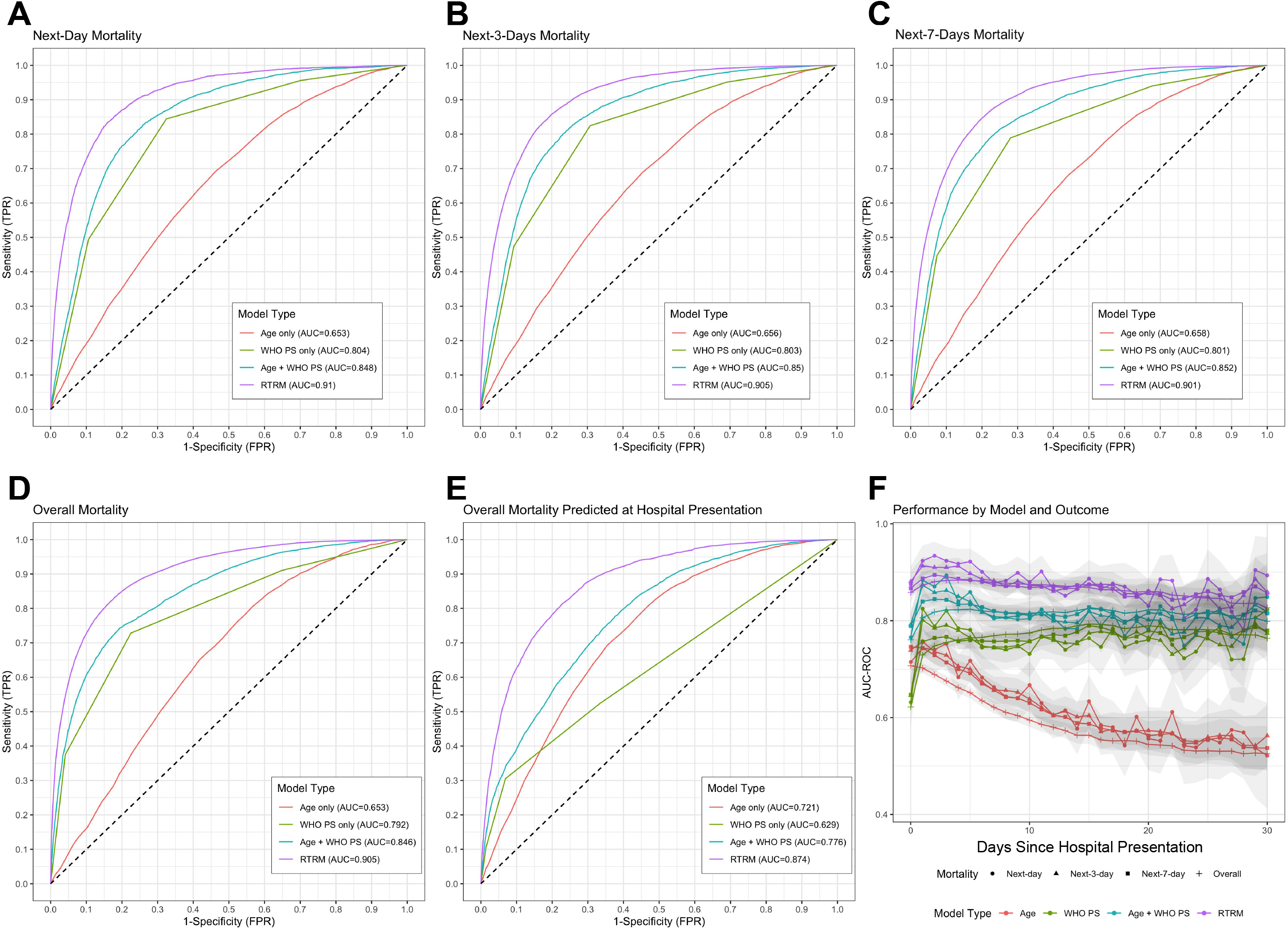
Receiver Operating Characteristic (ROC) Curves by Outcome and Model Type. Performance analysis of the RTRM measured by ROC, with models of the daily WHO Progression Scale (WHO PS) and Age as references. Performance is broken down by next-day, next-3-days, and next-7-days mortality, overall mortality, and mortality predicted at the date of presentation (A, B, C, D, and E respectively). Results are graphed by day since presentation (F), highlighting strong predictive power of the RTRM throughout COVID-19 hospitalization.

Using only baseline risk for prediction of overall mortality at hospital presentation rather than daily risk assessments, the RTRM achieved an AUCROC of 0.874, compared to 0.721, 0.629, and 0.776 for age and daily WHO PS alone and in combination, respectively (all *p*<0.0001; Figure 2E). Furthermore, the fact that the RTRM AUCROC of overall mortality using daily risk was 0.031 higher than the overall mortality at hospital presentation highlights the important contribution of longitudinal risk to model accuracy. While the baseline accuracy is high, the added value of the RTRM persists with a greater than 0.80 AUCROC over the entire course of hospitalization, out to 30 days (Figure 2F).

### RTRM daily risk predictions for next-day, next-3-days, next-7-days and overall mortality

A primary purpose of the RTRM is a more granular elucidation of deteriorating health and increased mortality risk for patients over their hospitalization. The performance of the RTRM is demonstrated quantitatively with AUCROCs for next-day, next-3-days, next-7-days, and overall mortality (Figure 2A-F) and by swimmer plots provided as visualization of individual patient risk across length-of-stay. Patients were grouped by their eventual outcome, death or discharge, to highlight the divergence in risk scores that was observable with the RTRM well in advance of their eventual outcome (Figure 3).

**Figure 3:**
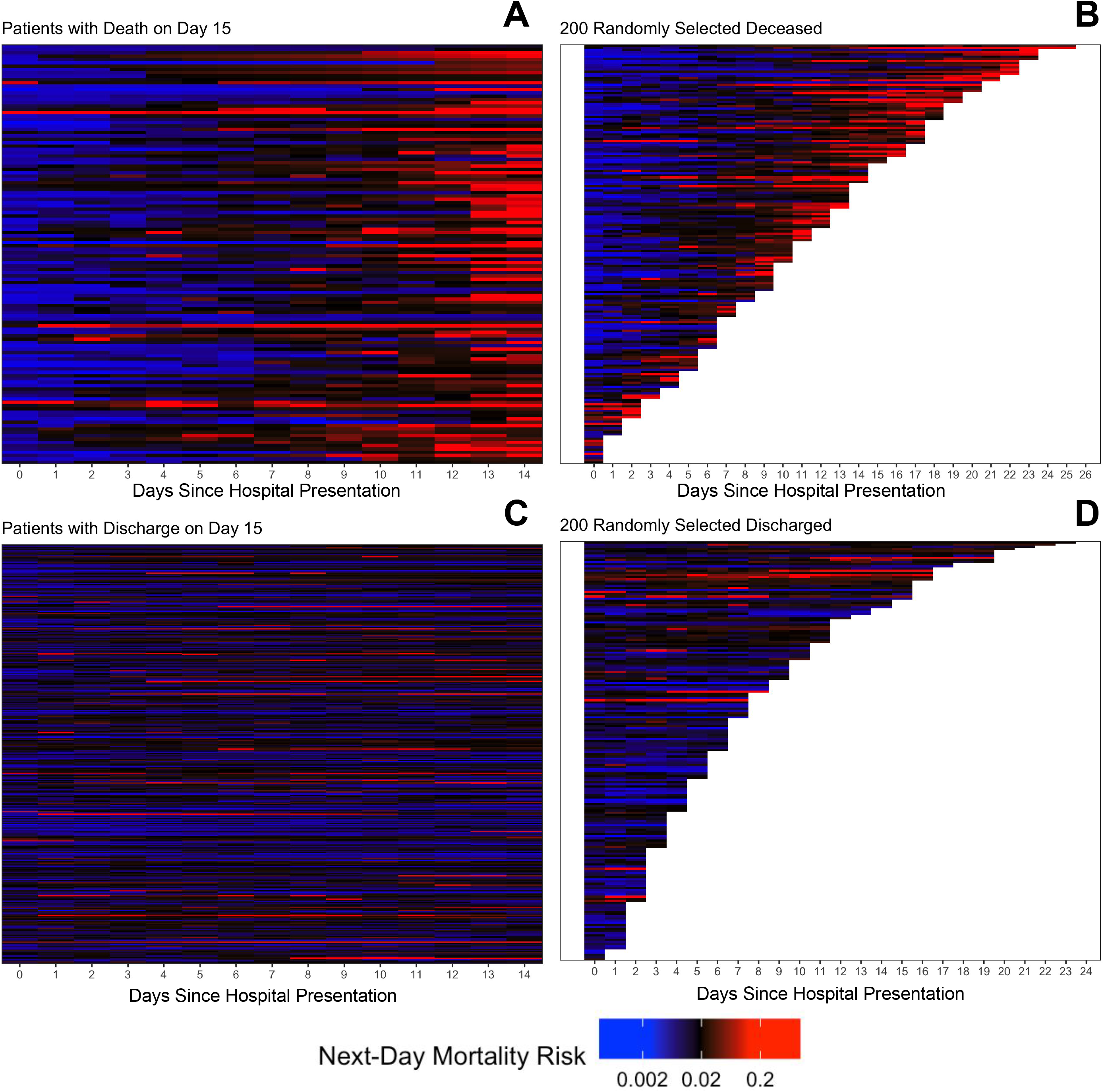
Swimmers Plots. RTRM scores (colored) for individual patients over time, selected based on eventual outcome. Patients were filtered at random from the test set for visualization purposes. Red cells indicate a higher risk of overall mortality and all patients experienced death (A, B) or discharge (C, D) at the end of their segments. Patients in A and C include only those that died or were discharged, respectively, on exactly the 15th day after hospital presentation, while B and D depict randomly selected patients that died or were discharged, respectively, at any time. Patterns suggest that the RTRM is able to reliably identify patients several days prior to their eventual death.

### RTRM prognostic factors

The evaluation of the most impactful predictors obtained from fitting the RTRM confirm previously known risk factors in COVID-19 and suggest interesting avenues for further investigation.^9^ Figure 4A shows the top 100 predictors by odds ratio (OR) associated with mortality by univariable analysis. Figure 4B shows the influence of all features in the RTRM. Although medications were excluded from the RTRM, frequencies of the predominant COVID-19 medications is provided in Table S4. Cut-points for each laboratory measure percentile included in the RTRM are provided (Table S5).

**Figure 4:**
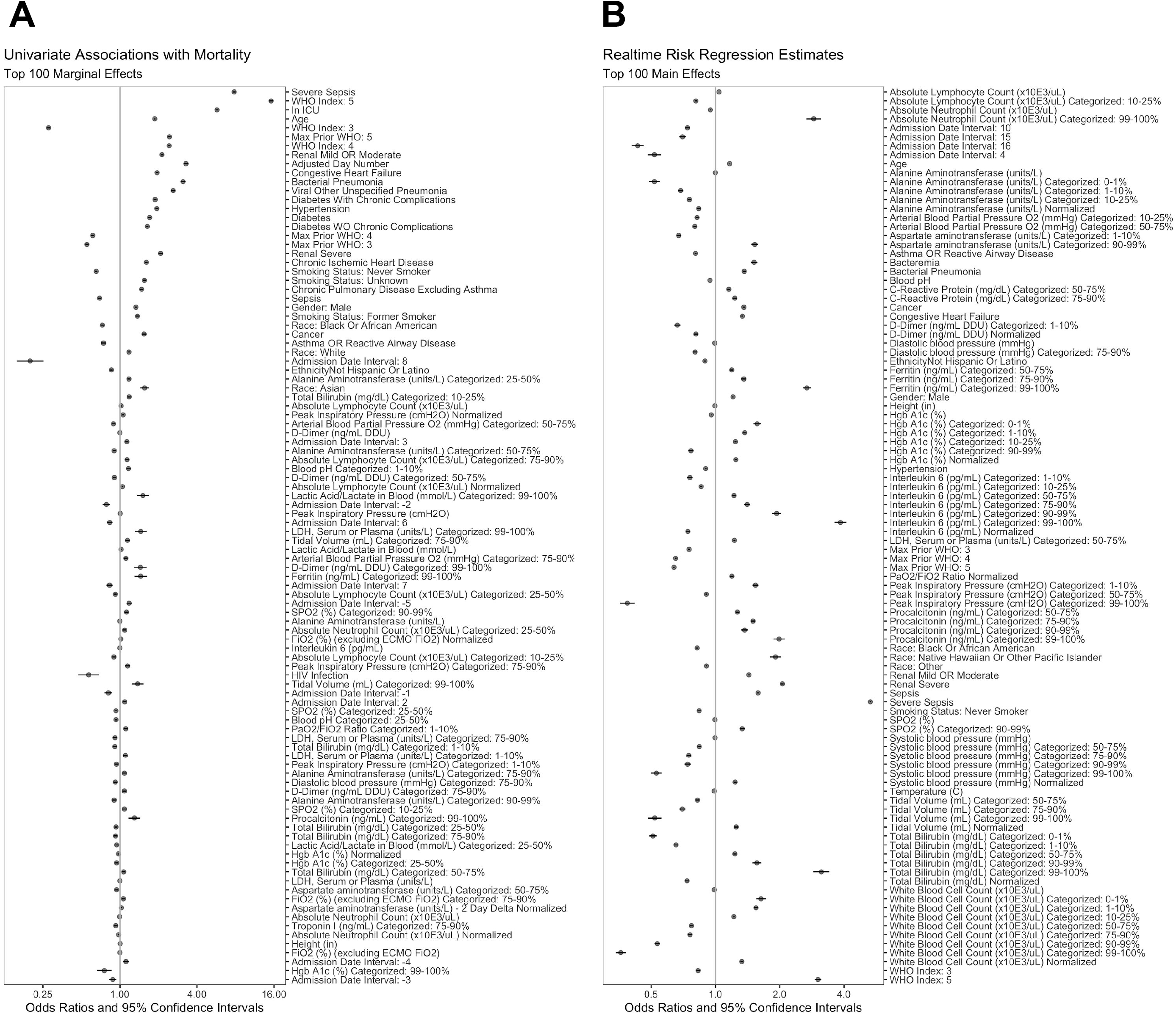
Forest Plots. Estimated parameters (odds ratio, OR) and 95% confidence intervals from the 100 most significant predictors of all-cause, hospitalized mortality based on either (A) univariable logistic regression or (B) multivariable logistic regression with elastic net (i.e. RTRM). Max Prior WHO and WHO Index are all referring to the WHO PS. ORs for categorical data are reported as normal for each categorical level independently. ORs for continuous variables are reported as the OR for a one standard deviation increase in the variable value.

Severe sepsis (OR=5.32, *p*<0.0001) and WHO PS of 5 (OR=3.02, *p*<0.0001) contribute the most to identifying future mortality (Figure 4). Age provides the greatest baseline demographic impact, with a 16.8 year increase in age associated with a 16.6% increase in mortality (*p*<0.0001); an effect likely minimized due to inclusion of competing risk factors such as comorbidities. The unadjusted association of age with mortality was found to be 1.87, 95% CI [1.84, 1.89] (Figure 4A). Figure S7 displays all variables used in the RTRM with the adjusted odds ratios. Overall, most of the blood markers showed appropriate dose responses with ferritin, interleukin-6, bilirubin, and white blood cell count having the largest distribution of response or ability to discriminate risk. Additionally, most comorbidities evaluated were significant independent predictors of mortality; albeit, the interpretation of the direction and effect size is limited due to the complexity of the RTRM (Figure S7).

### RTRM and calendar-time dependent risk

The mortality rate among patients hospitalized with COVID-19 has dropped substantially since the start of the pandemic.^10,11^ Improvements in clinical support care, changes in therapeutic regimens, evolution of the hospitalized patient population in regards to clinico-demographics, and alterations in hospital admission patterns or patient behavior have been speculated as reasons for improved mortality. The RTRM was utilized to investigate patterns across six epochs of time delineating major shifts in medication recommendations (Table 1). We evaluated the daily risk scores for all patients in our test set during hospitalization (Figure 5A, C) as well as restricted the evaluation to those patients who had not reached an endpoint for each day following hospital presentation (active; Figure 5B, D).

**Figure 5:**
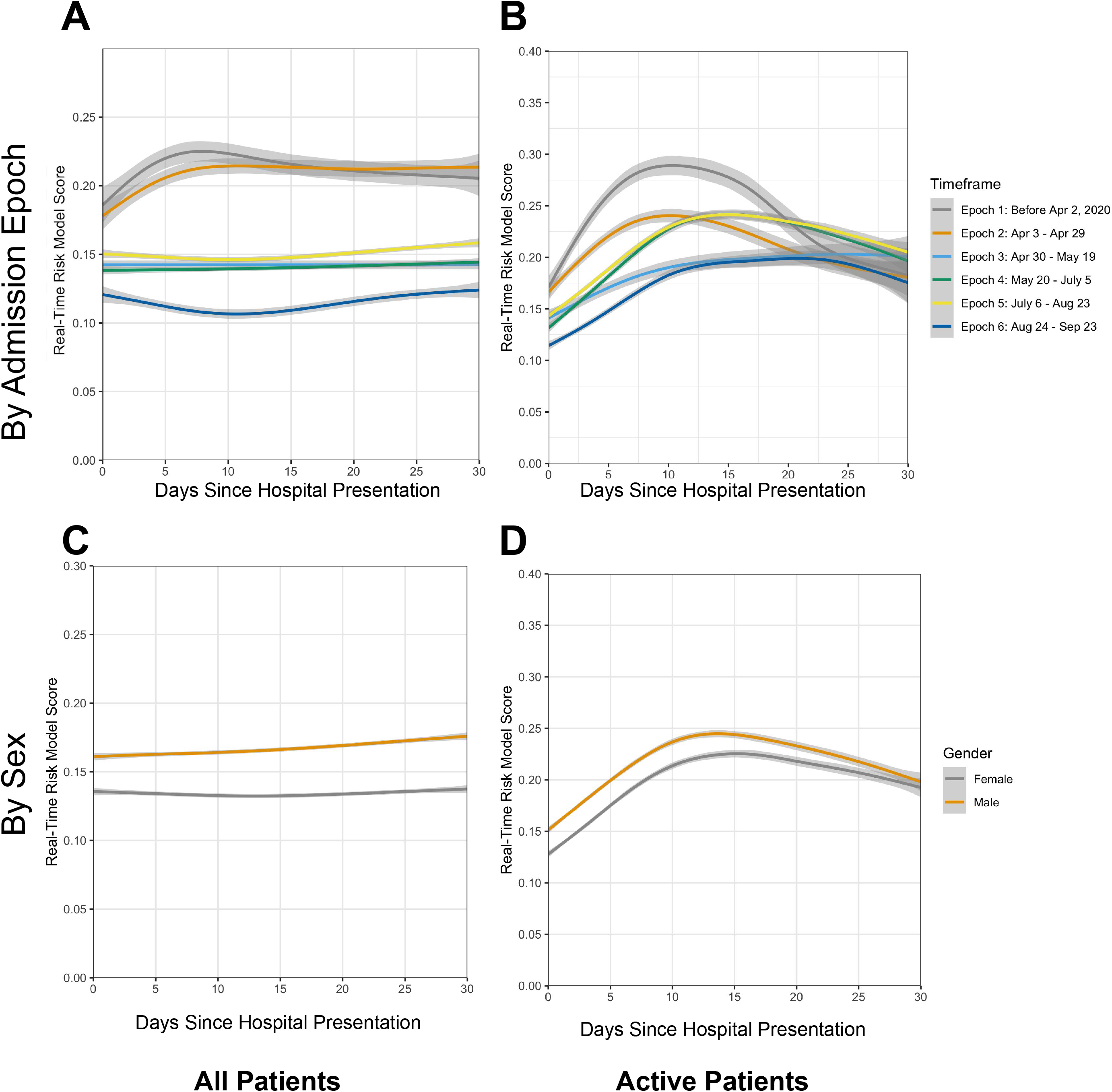
Real-Time Risk Curves. Smoothed and combined RTRM curves showing risk trajectories across six epochs and sex. A generalized additive model with integrated smoothness estimation was applied to the risk predictions over hospitalization time. Each point, *n*, on the y-axis represents the smoothed mean risk score for patients that were still hospitalized on the *n*^th^ day following COVID-19 hospital presentation. Patients that have experienced an outcome continue to contribute to the risk of mortality after their death or discharge in A and C; whereas, in B and D only active patients (i.e. still-hospitalized) contribute to the model for any given day after hospital presentation. RTRM curves are broken down by six calendar date epochs (A, B) and sex (C, D).

Figure 5A shows the RTRM risk over days from presentation and stratified by the six epochs. The patient cohort presenting to hospitals during Epochs 1 and 2 had substantially increased mortality compared to all others. Risk increased rapidly over the first 10 days of Epochs 1 (slope=0.0044, p<0.0001) and 2 (slope=0.0043, p<0.0001); whereas, Epochs 3 to 5 experienced no change in risk (Figure 5A). Strikingly, Epoch 6 actually experienced reductions in risk over the first 10 days (slope= -0.0019, p<0.0001). These reductions in slopes across epochs suggest improvements in care led to improved outcomes over time, marked by reductions in severe sepsis for example (Table 3). These data suggest improvement in clinical care may have occurred between Epochs 2 and 3, which corresponds with the shift in treatment recommendation towards the use of low-dose steroids in the late pulmonary phase, and between Epochs 5 and 6, which corresponds with a shift from restricting convalescent plasma to severe patients to less severe patients based on regulatory approvals (Table 1).

Furthermore, there was a meaningful separation of risk at hospital presentation (Day 0), with Epochs 1 and 2 at greatest risk, Epochs 3 to 5 having moderate risk, and Epoch 6 having much lower risk (Figure 5). This suggests that disease severity and/or risk factors evolved over time. Interestingly, neither age nor any of the comorbidities consistently changed over epochs. There were, however, relatively fewer Asian and African Americans and an increase in Caucasians by Epoch (Table 2). There was a negative trend in the frequency of baseline mechanical ventilation/ECMO with a concomitant increase in the use of non-invasive ventilation or high-flow oxygen at hospital presentation (Table 2). This suggests decreases in mortality rates over time may also be attributed to patients either presenting to the hospital or being admitted with less severe disease.

Lastly, showing the trajectory of RTRM mortality risk from days of hospital presentation for active patients only, indicate that risk of death increased for each day of hospitalization until days 10 to 14 (Figure 5B, D). Risk of mortality then remained stable or decreased, as visualized most strikingly in Epoch 1 but demonstrated by all other Epochs (Figure 5B) and both males and females (Figure 5D).

### Sex disparities in the RTRM

We evaluated the trajectories of risk scores according to sex. The smoothed risk averages displayed in Figure 5C and D suggest that disparities in sex mortality can be attributed to differences in risk at hospital presentation. Following the separation at day 0, both men and women follow similar risk trajectories for the first 20 days for both overall (Figure 5C) and active patients only (Figure 5D).

## Discussion

Quantifying the risk of mortality at a specified time for patients hospitalized with COVID-19 is critical to understanding the impact of clinical interventions. The widespread impact of the COVID-19 pandemic yielded a broad spectrum of patients with diversity across geography, timeframe, medical history, and demographics. Patient prognoses differ substantially based on these factors. Recent studies highlighted the importance of age and oxygen saturation as key prognostic factors by multivariable analyses, but none evaluated risk beyond admission.^5-8,12^

Utilizing EHRs for 46,971 patients hospitalized with COVID-19, we evaluated hundreds of elements to develop a real-time risk model (RTRM) of all-cause, hospitalized mortality. Both the daily WHO PS and the age models perform well at predicting mortality as previously reported.^4-6^ Therefore, it is noteworthy that the RTRM is superior to the combined age and the daily WHO PS model for next-day, next-3-days, next-7-days, and overall mortality. Additionally, due to longitudinal daily assessment, the RTRM was able to separately evaluate differences in mortality based on risk at presentation versus risk during the course of care. The RTRM revealed that reduction in mortality rates over time during the pandemic was associated with not only reduction in baseline risk due to disease severity and shifts in demographics, but also suggested improvement in care over time. Notably, we did not find differences in age across epochs. Although other studies have shown that males are at increased risk of death due to COVID-19, the RTRM suggests this increase is primarily due to severity at presentation rather than decreased response to clinical care. Finally, evaluation of effect size and significance of all predictors in the RTRM multivariable regression model not only corroborates most of the current key predictors of mortality in COVID-19 such as age, WHO PS, male sex, comorbidities, and markers of inflammation and organ dysfunction, but also shows clear dose responses for critical laboratory measures.^13^

Our study has several limitations, some of which is inherent to real-world EHR data. In particular, because ICU care status was based on EHR location data, ICU misclassification may have occurred due to creation of COVID-19 isolation wards within ICUs, regardless of the necessity for ICU-level care. Additionally, the complexity of the RTRM reduces its current applicability to clinical trials and other healthcare systems; however, we have provided a full list of variables and their contributions to prediction of hospitalized mortality, for further scrutiny and to allow incorporation of top contributors in other research and clinical applications.

Future directions could focus on both research and clinical applications. First, the depth of the cross-facility, harmonized EHR data allows for more robust risk adjustment in retrospective, matched cohort studies and provides a granular endpoint to evaluate interventions beyond general effectiveness towards optimal delivery, including timing and dosing. Indeed, medications were purposely excluded from the RTRM in order to facilitate the use of the model in downstream interventional studies. In an environment where randomized control trials are challenging to operate and recruitment is limited, the RTRM allows real-world evidence to identify which populations benefit from an intervention or combination of interventions. Second, based on the top contributors to the RTRM that can be reliably measured, a simplified risk score can be developed to provide ease of calculation and broader applicability. In addition, RTRMs that predict other clinical outcomes, such as progression of pulmonary disease and development of sepsis and/or multiorgan failure should be investigated. Future work should consider readmissions related to COVID-19 complications and chronic COVID-19 etiologies, which is particularly relevant given studies show that 9-15% of patients hospitalized due to COVID-19 and discharged are readmitted within 60 days.^14,15^ In summary, to our knowledge, the RTRM is the first of its kind to evaluate daily risk of hospitalized mortality for patients with COVID-19. Not only does the RTRM outperform benchmark predictors of mortality, but it lays down a framework for optimizing future research and personalized care utilizing real-world evidence.

## Data Availability

The data that support the findings of this study are available upon request from the corresponding author. The data are not publicly available due to privacy restrictions. Extensive detail on all variables utilized by the RTRM has been provided in the supplemental materials, including significance and effect size.

## Contributors

DS contributed to study design, all analyses, modeling, results interpretation and manuscript preparation. AMF contributed to data curation, sourcing and mapping as well as manuscript preparation and review. JC contributed to data mapping, data verification, manuscript preparation and review. XF contributed to data mapping, data verification, and analysis review. CM contributed to EHR data curation and sourcing, data verification, and manuscript review and editing. AJ contributed to study design, data verification and manuscript review and editing. HAB provided general oversight and contributed to results interpretation and manuscript review. LES provided oversight and contributed to overall study design, results interpretation, and manuscript review. GF provided general oversight and contributed to results interpretation and manuscript review. MC contributed to overall study design, data acquisition, and results interpretation. CFL contributed to study design, results interpretation and manuscript review. SAAE oversaw all aspects of the study including conceptualizing the study design and directing its implementation, as well as contributed to the development of the statistical analysis plan, interpretation of results, and manuscript writing.

## Declaration of Interests

The views expressed in this publication represent those of the authors and do not necessarily represent the official views of HCA Healthcare or any of its affiliated entities. None of the authors declared any conflict of interest related to the current study beyond employment with an affiliate of HCA Healthcare. HAB reported grant funding from MedImmune, Boehringer Ingelheim, Merck, Moderna, Verastem, Harpoon, Jounce, Janssen, BIND Therapeutics, Pfizer, Vertex, Gilead, Bayer, Incyte, AstraZeneca, Novartis, Seattle Genetics, GlaxoSmithKline, BioAtla, Agios, BioMed Valley, TG Therapeutics, eFFECTOR, CicloMed, Array, Roche/Genentech, Arvinas, Bristol-Myers Squibb, Macrogenics, CytomX, Arch, Revolution Medicine, Lilly, Tesaro, Takeda/Millennium, miRNA, Kyocera and Foundation Medicine (all paid to his institution); consulting fees from Incyte, AstraZeneca, Celgene, and Forma Therapeutics (all paid to his institution); non-compensated consulting services from Novartis, Bayer, Pfizer, GRAIL and Daiichi Sankyo; expert testimony from Novartis (paid to his institution) and stock ownership in HCA Healthcare. DS, AMF, JC, MC, and SAAE reported stock ownership in HCA Healthcare.

## Acknowledgements

The authors wish to acknowledge Drs. Susan Garwood, Marjorie Wongskhaluang, Joe Restivo, and Faisal Cheema for their clinical insight as well as Troy Gifford, Ryan Patrick, Cecile Roman, Daniel Luckett, and Shaita Picard for their contributions to electronic data extraction, management and curation. Thank you to Molly Altman for providing project management and general manuscript support. We would also like to acknowledge Mandelin Cooper, Mitch Davis and Laura McLean in the Clinical Operations Group for their parallel COVID-19 work that provided insight into HCA Healthcare treatment recommendations for the date epochs as well as variable sourcing and mapping suggestions.

## Supplemental Materials

### Supplemental Methods

#### Data Generation and Cleanup

Data was obtained from Genospace, an internal platform at HCA Healthcare that allows live querying of data obtained from HCA Healthcare’s Electronic Data Warehouse.

All preprocessing was performed with the intention of generating a patient-day matrix where each row represented the available information for an individual patient on an individual day. The columns of this matrix contain all available data for that patient-day, including demographics, labs, vitals, oxygenation, and hospitalization status. Many variables are not necessarily available on the single-day level. For variables not associated with a specific date in the dataset, such as race, age, or weight, the values were repeated for each patient-day of hospitalization. Other values, such as vitals, were commonly available with much greater than single-day frequency. For any such variable containing multiple instances in a single day, the mean for each patient-day was taken. The minimum and maximum were also considered, but were not found to improve model performance on the whole due to added model complexity. This process was applied for all continuous variables. For WHO PS, which is based on the level of respiratory support received, the maximum value, representing the most severe level of respiratory support, for a given day was used rather than the mean, except if the patient was discharged alive, in which case a score of 1 was assigned. For categorical values with dates, such as comorbidities and complications, including pneumonia, sepsis, and bacteremia, the indication of the event was added to the given patient-day and all subsequent days.

Missing value handling depended on the variable (Figure S1). For multi-level factors, e.g. race, patients with missing values were specifically given the value “Missing”. For binary indicator variables, e.g. comorbidities, the absence of a data point for a particular patient was treated as a negative. For continuous values, e.g. lab results, data was completed with multiple imputation, as described in the main text and below in the Imputation Evaluation section.

#### Lambda Hyperparameter Tuning

The Real-Time Risk Model requires selection of shrinkage coefficients to be estimated for L1 (absolute) and L2 (squared) regularization. This process was performed with 10-fold cross validation and a grid search where L1 and L2 were chosen independently. For each point in the grid, the risk model was fit for each fold and evaluated on the cross validation test set. The deviance of the model was measured at each stop. Hyperparameters were chosen based on the deviance minimization within the grid search.

#### Interactions Evaluation

The Real-Time Risk Model considered interactions between pairwise combinations of variables, i.e. the possible altering of the linear effect of a variable on mortality conditional on another variable. We allowed this complexity into our model with the introduction of simple interacting coefficients for selected variables. Given the large number of coefficients to be considered, we recognized that including all n-choose-2 pairwise combinations would dramatically increase the dimensionality of our model. To mitigate this, we limited the number of coefficients to those with higher clinical or statistical support. Interactions were modeled based on all pairwise combinations of the following ten variables - (1) Age, (2) WHO PS, (3) Admission Date, (4) ICU Status, (5) Hospitalization day number, (6) C-Reactive Protein (mg/dL), (7) D-Dimer (ng/mL DDU), (8) Systolic Blood Pressure (mmHg), (9) Temperature (C), and (10) SPO2 (%). These terms formed 45 pairwise interaction terms (Figure S4). The conditional distributions of each of the interacting variables were evaluated (Figure S6).

#### Imputation Evaluation

While missing data at any point after a baseline value can be directly inferred from the last available value, missing baseline data as well as values for variables not associated with a specific date in the dataset needed to be imputed from available data. To prevent forward-looking data leakage, no timepoint data imputation was permitted to utilize data that was not available at the time of the missing datapoint. To perform this imputation, we used Multiple Imputation by Chained Equations (MICE).^1^ With this process, missing data for each variable was randomly sampled with replacement from the observed data. Then, for each variable, missing values were removed and replaced with fitted values from regressing the observed values for that variable against the rest of the dataset. This step was performed for each variable containing any missing data and repeated until convergence. The stochastic component of this procedure was designed to capture some of the variability present in the original data. Consequently, we produced 10 separate imputed datasets in this manner and executed our analysis separately on each. The results of each analysis were merged by combining, via simple average, the resulting coefficients into a single prediction model.^2^

While all iterations of the multiple imputation method differed slightly from one another, the predicted probabilities were highly correlated (r = 0.977) and the merged model outperformed each individual model on its own (Figure S5).

Variance for coefficients was measured with bootstrapping. Models were fit, as described above, repeatedly on prediction matrices obtained by resampling the patient population with replacement. This process was repeated 100 times, with the standard deviation of the bootstrapped coefficient estimates taken as the bootstrap estimate of the standard errors, which are visualized and reported in Forest plots (Figure 4, Figure S6).

#### Model Fitting and Performance

Model validation was performed by applying the prediction coefficients generated from the training set to the held-back testing set. This entire process was performed ten times; once for each imputation step. Prediction scores for each imputation, corresponding to probability of *n*-day mortality, were averaged into a single *n*-day risk of mortality. Each patient-day was evaluated for the probability of *n*-day and overall survival. The model performance was measured with the area under the curve of the receiver operating characteristic (AUCROC). This analysis was performed using the R packages *ROCit, oem (Orthogonalizing EM: Penalized Regression for Big Tall Data)* and *glmnet*.^3-5^

---

**Figure S1:**
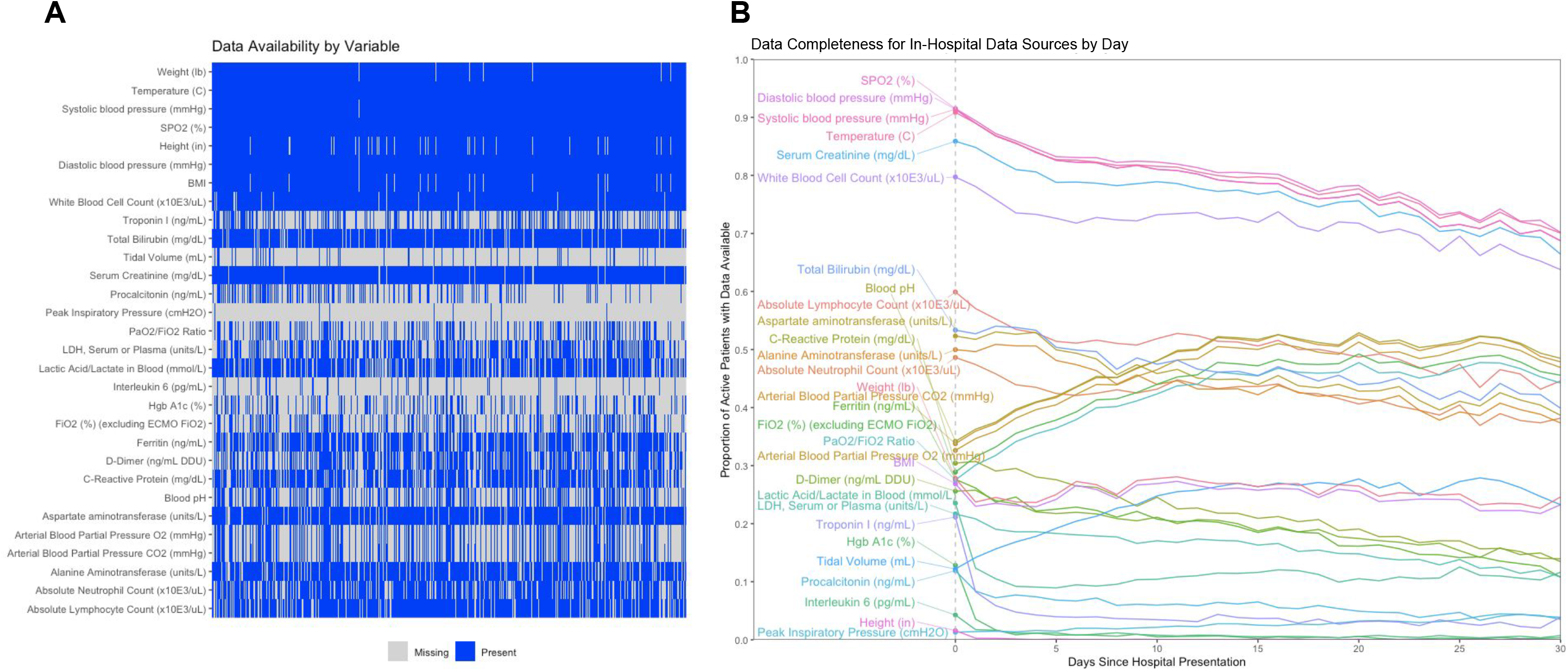
Data Completeness Summary. Summary of missing data used in the RTRM. (A) Describes the presence of any observed value (blue) present for an individual patient (x-axis) and key variable (y-axis). (B) Describes the proportion of observed values for key variables at individual points in time (x-axis).

**Figure S2:**
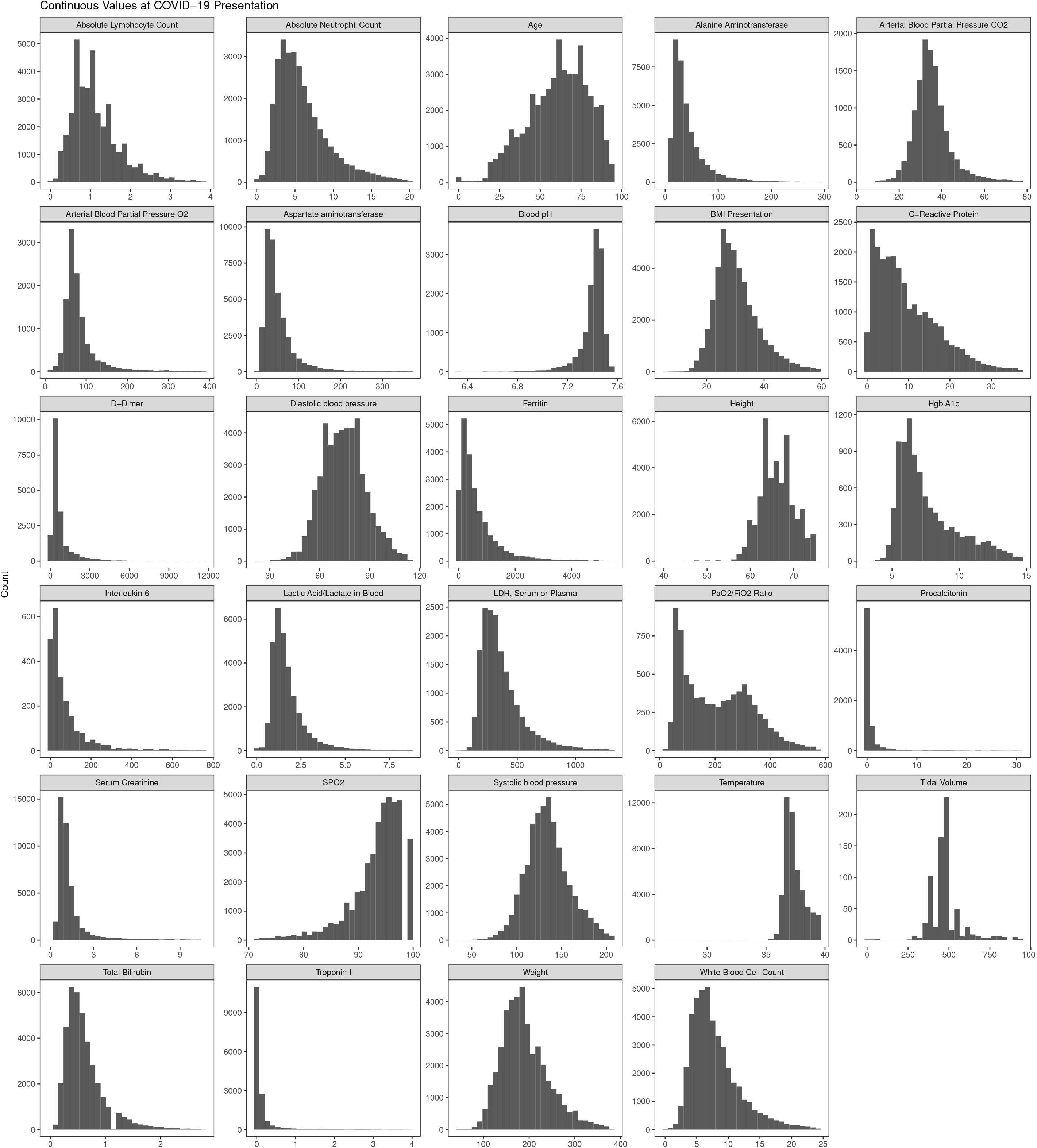
Continuous Data Variable Distributions. Distributions for raw values for continuous variables included in the RTRM.

**Figure S3:**
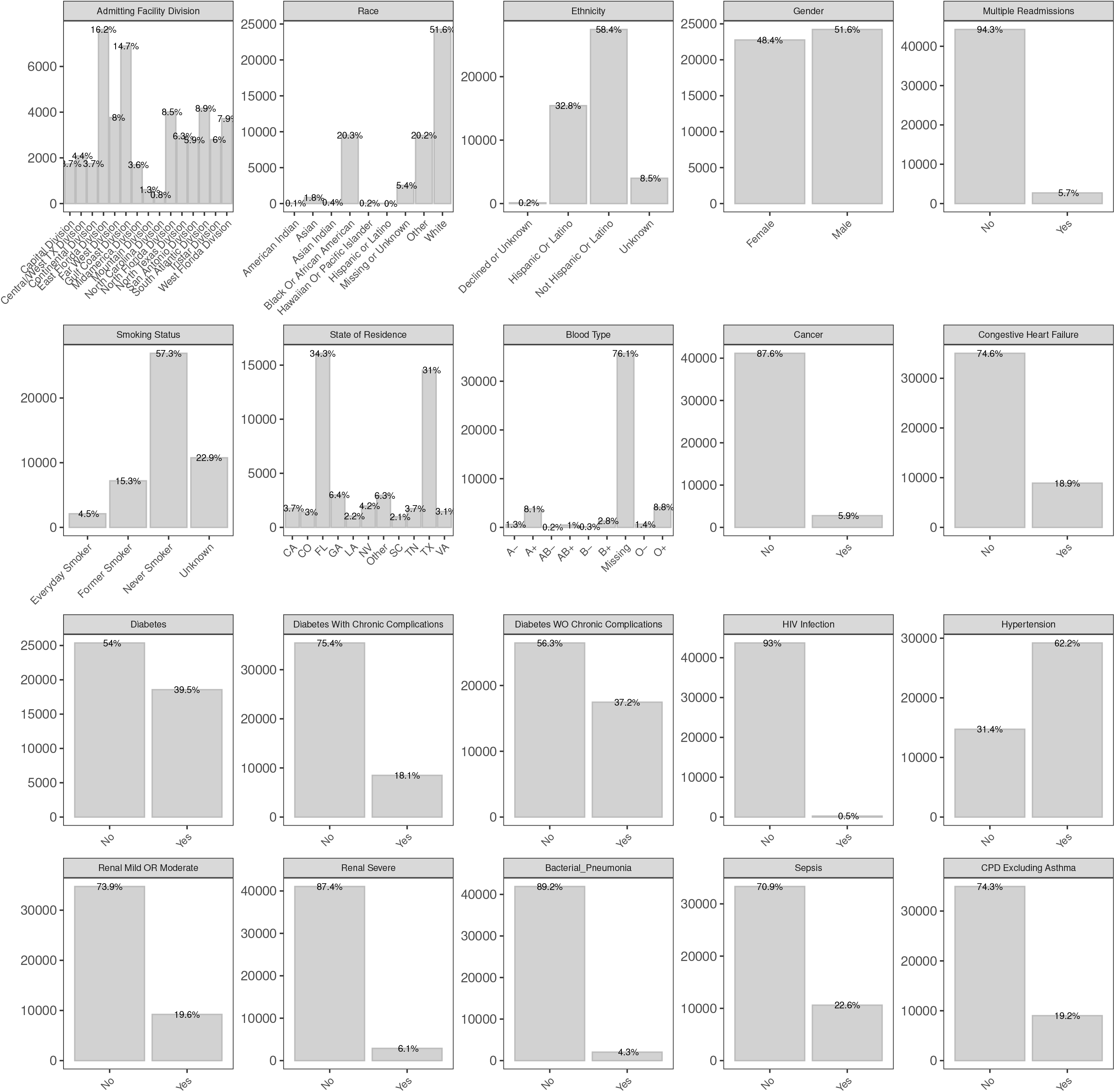
Categorical Data Variable Distributions. Distributions for raw values for categorical variables included in the RTRM.

**Figure S4:**
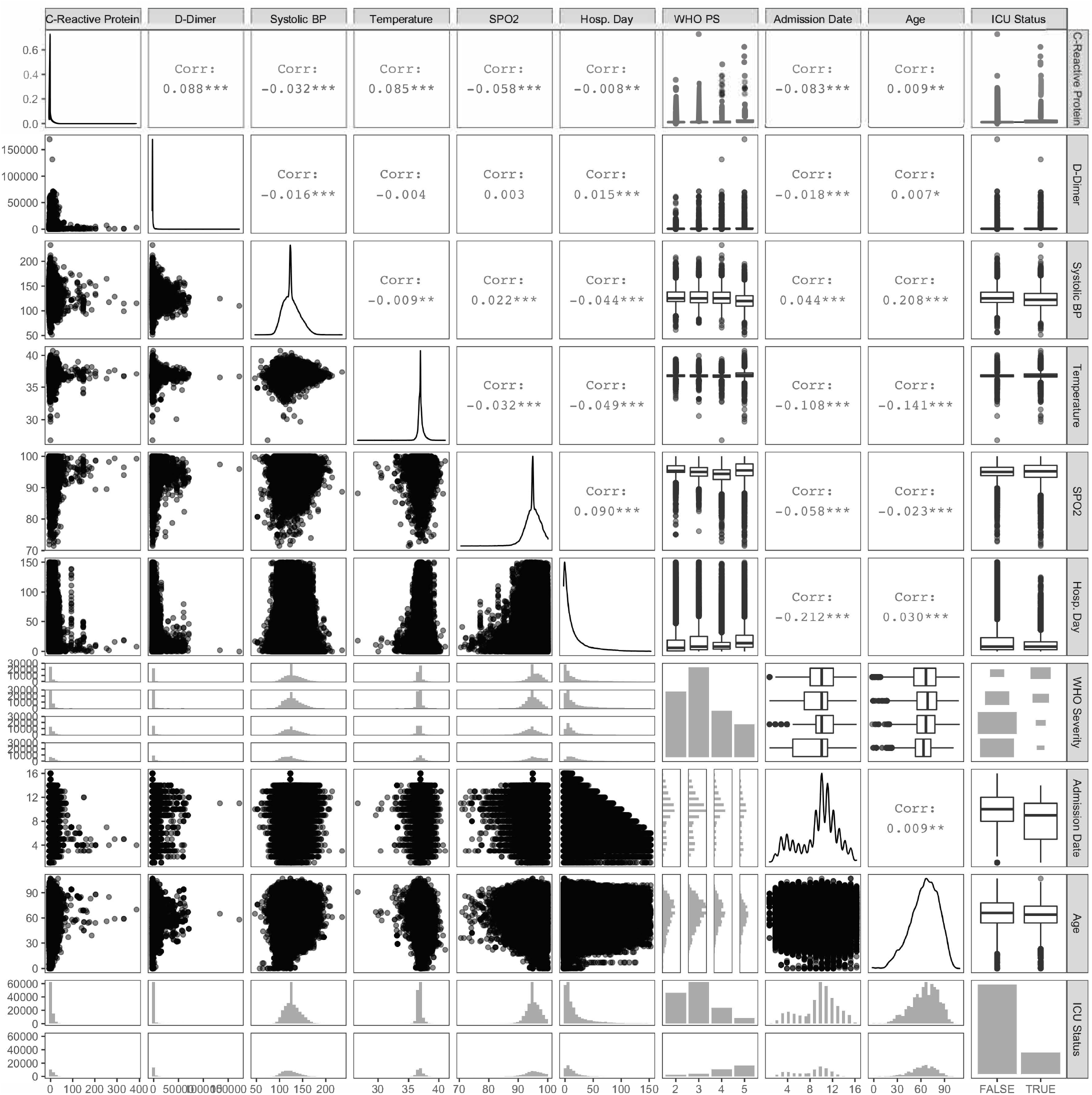
Interaction Pairwise Plots. Pairwise plots summarizing the bivariate distributions of all variables included as interaction terms in the RTRM.

**Figure S5:**
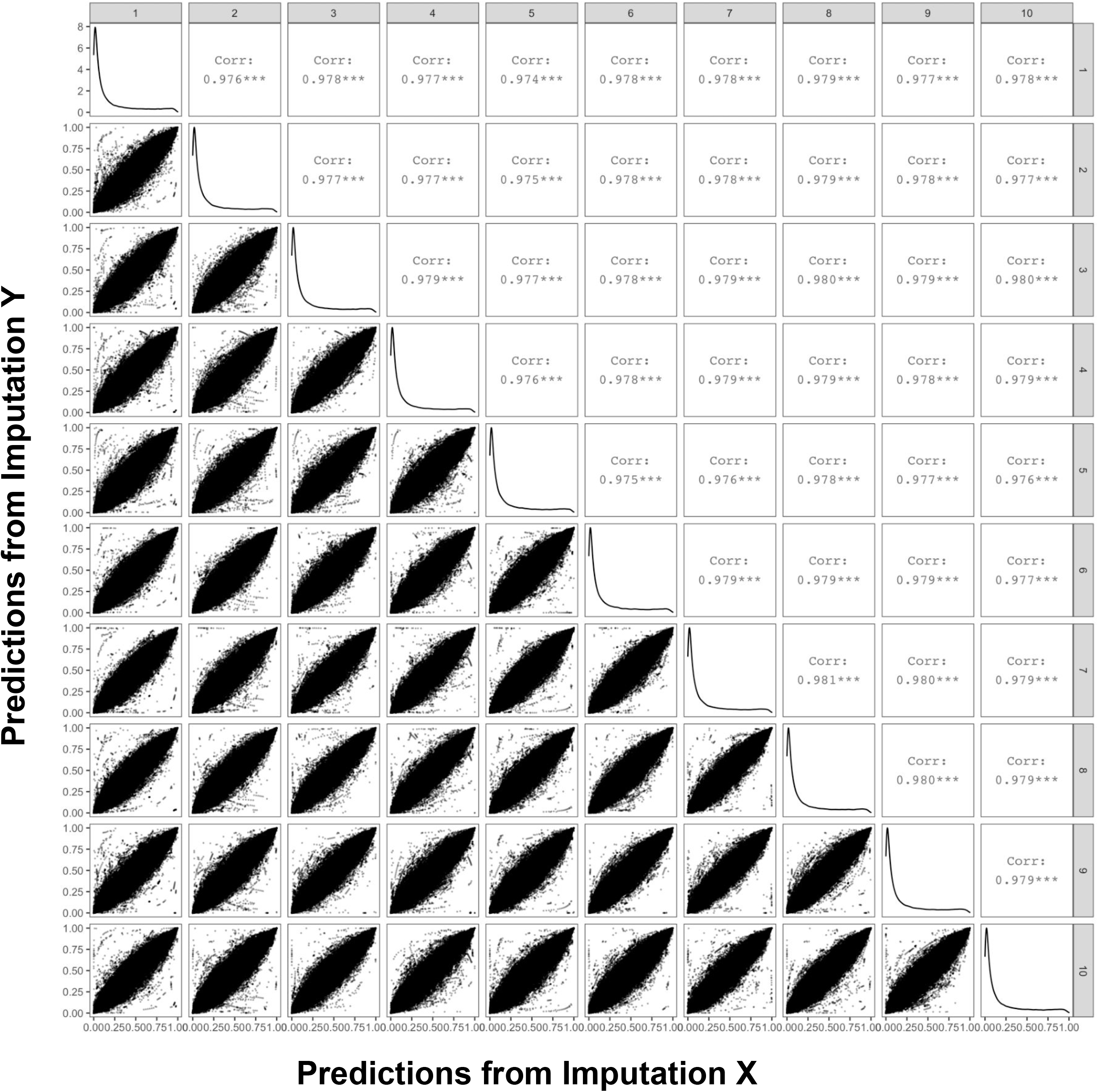
Imputation Pairwise Plots. Pairwise correlation scatterplots for RTRM scores derived from all ten individual imputations. Strong and consistent correlation between imputation runs suggest very little dependency on the randomization component for completing missing data.

**Figure S6:**
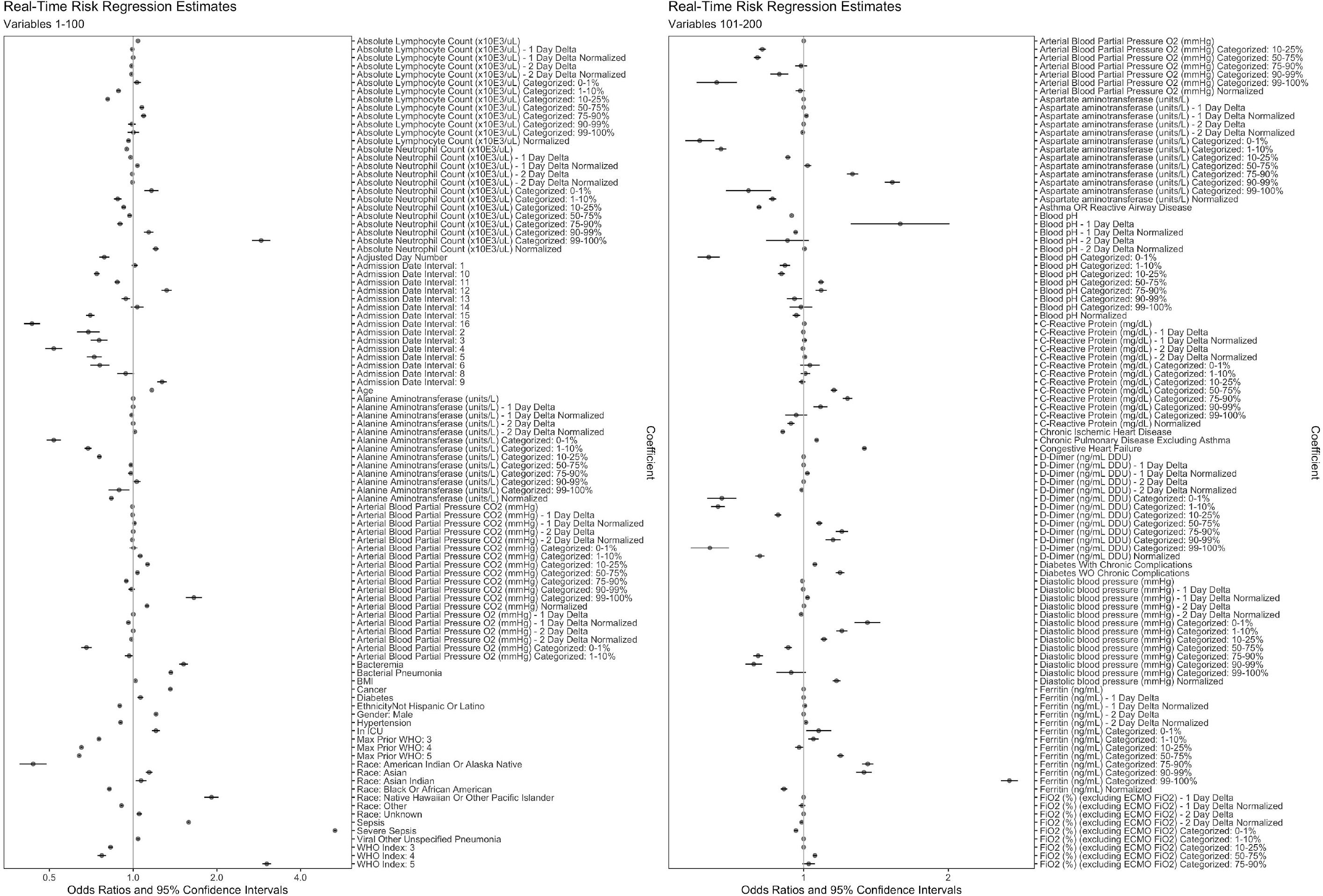
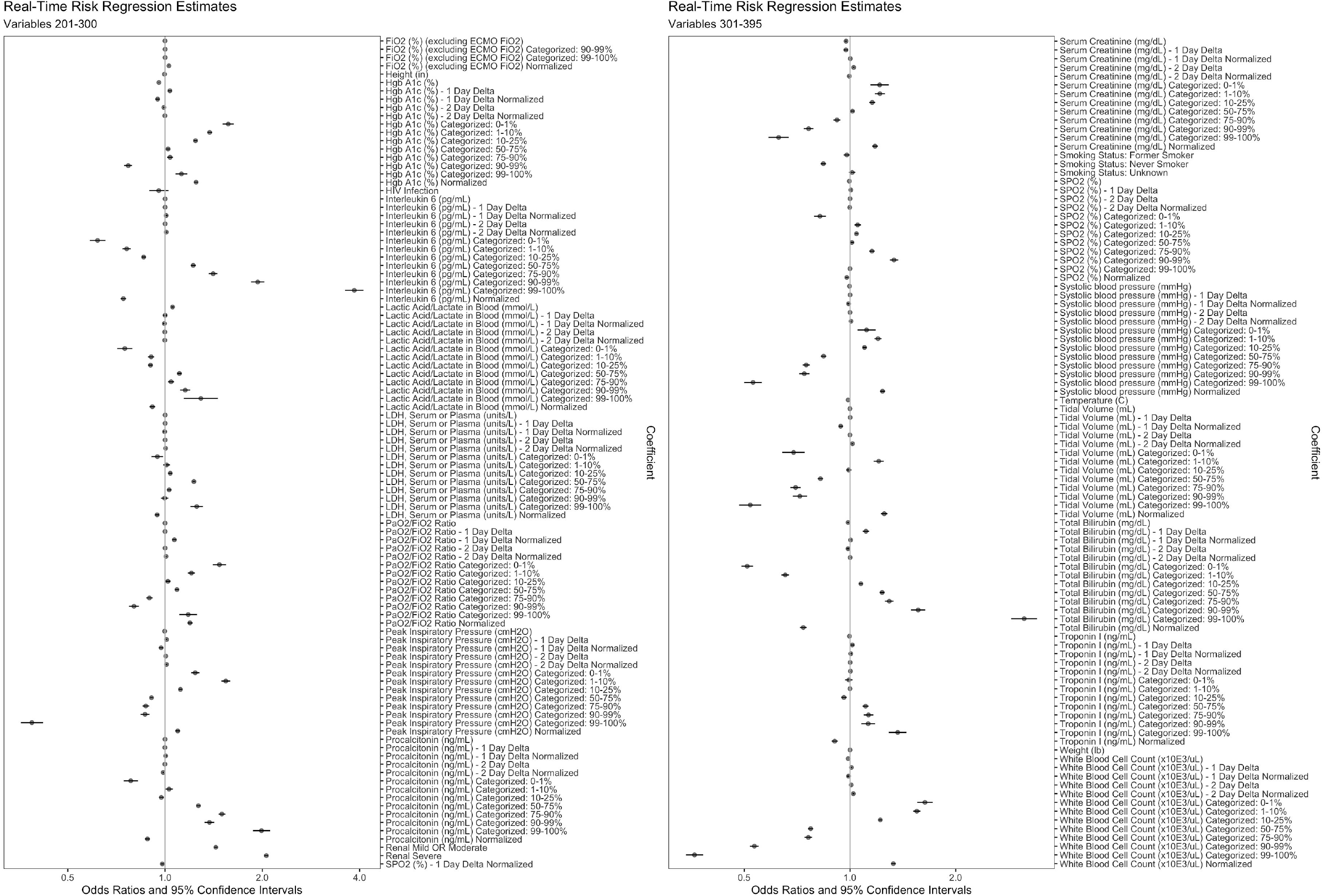
Forest Plots for All Variables in the RTRM. Estimated parameters (odds ratio, OR) and 95% confidence intervals from all features in the RTRM. Max Prior WHO and WHO Index are all referring to the WHO PS. ORs for categorical data are reported as normal for each categorical level independently. ORs for continuous variables are reported as the OR for a one standard deviation increase in the variable value.

## Supplemental Tables

**Table S1:** ICD-9-CM and ICD-10-CM codes for Identification of Comorbidities in Electronic Health Record Data.

| Comorbidity | ICD-9-CM | ICD-10-CM |
| --- | --- | --- |
| Hypertension | 401-405 | I10-I16 |
| Congestive heart failure* | 398.91, 402.01, 402.11, 402.91, 404.01, 404.03, 404.11, 404.13, 404.91, 404.93, 425.4-425.9, 428 | I11.0, I13.0, I13.2, I25.5, I42.0, I42.5-I42.9, I43, I50, P29.0 |
| Chronic ischemic heart disease | 412, 414 | I25 |
| Asthma or reactive airway disease* † | 493 | J45 |
| Chronic pulmonary disease (excluding asthma)* | 490-492, 494-496, 500-505, 506.4, 508.1, 508.8 | J40-J44, J47, J60-J67, J68.4, J70.1, J70.3 |
| Diabetes | 249.0-249.3, 249.9, 250 | E08-E13 |
| Diabetes without chronic complications* | 249.0-249.3, 249.9, 250.0-250.3, 250.7-250.9 | E08.0, E08.1, E08.6, E08.8, E08.9, E09.0, E09.1, E09.6, E09.8, E09.9, E10.1, E10.6, E10.8, E10.9, E11.0, E11.1, E11.6, E11.8, E11.9, E13.0, E13.1, E13.6, E13.8, E13.9 |
| Diabetes with chronic complications* | 250.4-250.7 | E08.2, E08.3, E08.4, E08.5, E09.2, E09.3, E09.4, E09.5, E10.2, E10.3, E10.4, E10.5, E11.2, E11.3, E11.4, E11.5, E13.2, E13.3, E13.4, E13.5 |
| Cancer* | 140-149, 150-159, 160-165, 170-172, 174-176, 179-189, 190-198, 199.0, 199.1, 200-208, 238.6 | C00-C14, C15-C26, C30-C39, C40-C41, C43, C45-C49, C50, C51-C58, C60-C63, C76-C80, C81-85, C88, C90-C96 |
| HIV infection* | 042 | B20 |
| Renal disease - mild or moderate* | 403.00, 403.10, 403.90, 404.00, 404.01, 404.10, 404.11, 404.90, 404.91, 582, 583, 585.1-585.4, 585.9, V42.0 | I12.9, I13.0, I13.10, N03, N05, N18.1-N18.4, N18.9, Z94.0 |
| Renal disease - severe*‡ | 403.01, 403.11, 403.91, 404.02, 404.03, 404.12, 404.13, 404.92, 404.93, 585.5, 585.6, 588.0, V45.1, V56 | I12.0, I13.11, I13.2, N18.5, N18.6, N25.0, Z49, Z99.2 |

\* The ICD-9 and ICD-10 codes used to identify these comorbidities were largely based on a modified Charlson Comorbidity Index (CCI) coding scheme (https://www.ncbi.nlm.nih.gov/pmc/articles/PMC6684052/)
† The diagnosis codes for asthma were separated from other chronic pulmonary diseases to facilitate the separate assessment of asthma, and because the same included diagnosis code can be used for both asthma and reactive airway disease, the label for this comorbidity is "Asthma or reactive airway disease"; the World Health Organization (WHO) ICD-10 code J46 was not included, as this code is not included in ICD-10-CM
‡ The diagnosis codes for unspecified renal failure (i.e., the ICD-9 code 586 and the ICD-10 code N19) were not included, as these codes are not sufficient to identify previous acute renal failure, which may have been reversible, vs. chronic kidney disease (e.g., end-stage renal disease) vs. acute renal failure/acute kidney injury as a complication of COVID-19

**Table S2:** ICD-10-CM codes for Identification of Pneumonia, Sepsis, and Bacteremia in Electronic Health Record Data*.

| Condition | ICD-10-CM |
| --- | --- |
| Bacteremia | R78.81 |
| Bacterial pneumonia | A01.03, A02.22, A37.01, A37.11, A37.81, A37.91, A54.84, J13-J15, J16.0, P23.1, P23.2, P23.3, P23.4, P23.5, P23.6 |
| Sepsis (excluding severe sepsis) | A02.1, A22.7, A26.7, A32.7, A40, A41, A42.7, B37.7, O85, O86.04, P36 |
| Severe sepsis | R65.2 |
| Viral/other/unspecified pneumonia | B01.2, B05.2, B06.81, B77.81, J09.X1, J10.0, J11.0, J12, J16.8, J17, J18, P23.0, P23.8, P23.9 |

\* Dates associated with these ICD-10 codes were filtered to align with COVID-19 diagnosis, including only dates in the range of two weeks before the collection date of the first positive SARS-CoV-2 test to 90 days after the first encounter with SARS-CoV-2 infection

**Table S3:** Pairwise Testing of Next n-Day and Overall Mortality AUCROCs*.

| Model | Mortality Type | Age | WHO PS | Age + WHO PS | RTRM |
| --- | --- | --- | --- | --- | --- |
| Age | Next-day | 1 | <0.00001 | <0.00001 | <0.00001 |
| WHO PS |  | <0.00001 | 1 | <0.00001 | <0.00001 |
| Age + WHO PS |  | <0.00001 | <0.00001 | 1 | <0.00001 |
| RTRM |  | <0.00001 | <0.00001 | <0.00001 | <0.00001 |
| Age | Next-3-day | 1 | <0.00001 | <0.00001 | <0.00001 |
| WHO PS |  | <0.00001 | 1 | <0.00001 | <0.00001 |
| Age + WHO PS |  | <0.00001 | <0.00001 | 1 | <0.00001 |
| RTRM |  | <0.00001 | <0.00001 | <0.00001 | 1 |
| Age | Next-7-day | 1 | <0.00001 | <0.00001 | <0.00001 |
| WHO PS |  | <0.00001 | 1 | <0.00001 | <0.00001 |
| Age + WHO PS |  | <0.00001 | <0.00001 | 1 | <0.00001 |
| RTRM |  | <0.00001 | <0.00001 | <0.00001 | 1 |
| Age | Overall | 1 | <0.00001 | <0.00001 | <0.00001 |
| WHO PS |  | <0.00001 | 1 | <0.00001 | <0.00001 |
| Age + WHO PS |  | <0.00001 | <0.00001 | 1 | <0.00001 |
| RTRM |  | <0.00001 | <0.00001 | <0.00001 | 1 |

\* *p*-values are reported.

**Table S4:**
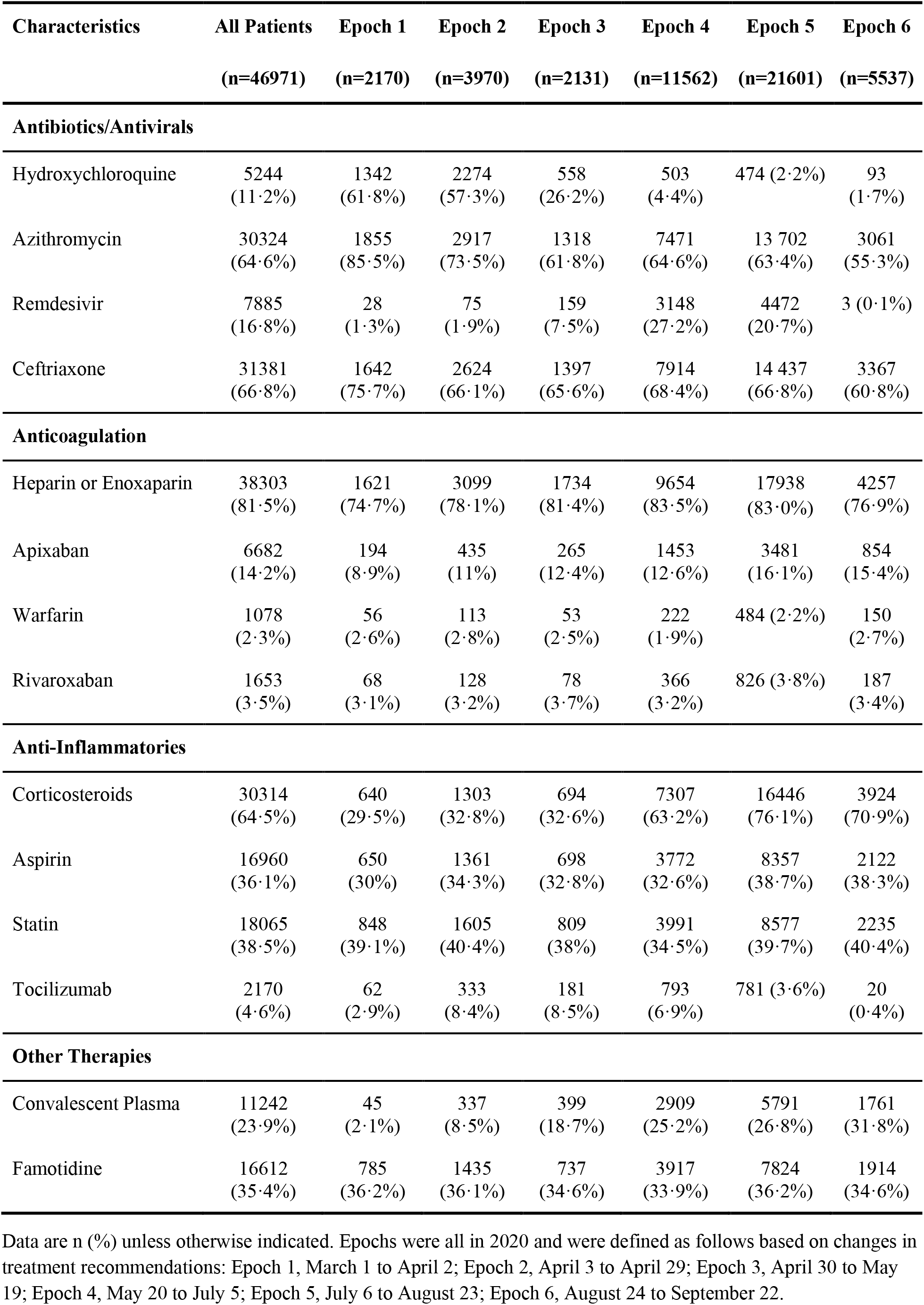
Medications.

**Table S5:** Laboratory and Vital Values by Patient Percentiles.

| Lab or Vital Test | 1% | 10% | 25% | 50% | 75% | 90% | 99% |
| --- | --- | --- | --- | --- | --- | --- | --- |
| Absolute Lymphocyte Count (x10E3/uL) | 0.11 | 0.40 | 0.60 | 0.97 | 1.40 | 2.00 | 3.85 |
| Absolute Neutrophil Count (x10E3/uL) | 1.2 | 2.9 | 4.5 | 7.1 | 10.7 | 15.0 | 26.3 |
| Alanine Aminotransferase (units/L) | 9 | 16 | 24 | 38 | 65 | 117 | 675 |
| Arterial Blood Partial Pressure CO2 (mmHg) | 22.3 | 30.0 | 34.6 | 40.9 | 50.0 | 61.8 | 91.5 |
| Arterial Blood Partial Pressure O2 (mmHg) | 41.0 | 54.9 | 63.1 | 75.9 | 97.2 | 136.2 | 275.7 |
| Aspartate aminotransferase (units/L) | 10 | 18 | 26 | 39 | 61 | 101 | 647 |
| Blood pH | 7.06 | 7.26 | 7.34 | 7.41 | 7.45 | 7.49 | 7.55 |
| C-Reactive Protein (mg/dL) | 0.4 | 1.1 | 2.5 | 6.0 | 12.5 | 19.4 | 35.8 |
| D-Dimer (ng/mL DDU) | 125 | 230 | 355 | 650 | 1496 | 3375 | 15100 |
| Ferritin (ng/mL) | 24 | 125 | 273 | 548 | 1020 | 1819 | 6825 |
| FiO2 (%) (excluding ECMO FiO2) | 21 | 35 | 50 | 70 | 100 | 100 | 100 |
| Hgb A1c (%) | 4.8 | 5.5 | 6.0 | 6.9 | 8.8 | 11.3 | 14.6 |
| Interleukin 6 (pg/mL) | 1.5 | 6.2 | 16.9 | 48.5 | 129.3 | 365.9 | 2033.7 |
| Lactic Acid/Lactate in Blood (mmol/L) | 0.5 | 0.9 | 1.1 | 1.5 | 2.1 | 3.1 | 11.4 |
| LDH, Serum or Plasma (units/L) | 138 | 204 | 261 | 354 | 499 | 714 | 1753 |
| PaO2/FiO2 Ratio | 44.4 | 61.8 | 81.6 | 123.4 | 191.6 | 282.1 | 460.9 |
| Peak Inspiratory Pressure (cmH2O) | 10 | 14 | 18 | 24 | 30 | 36 | 47 |
| Procalcitonin (ng/mL) | 0.03 | 0.06 | 0.11 | 0.28 | 1.06 | 4.55 | 48.80 |
| Serum Creatinine (mg/dL) | 0.30 | 0.52 | 0.70 | 0.90 | 1.41 | 3.10 | 9.53 |
| Tidal Volume (mL) | 174 | 350 | 400 | 450 | 500 | 530 | 748 |
| Total Bilirubin (mg/dL) | 0.2 | 0.3 | 0.4 | 0.5 | 0.8 | 1.1 | 4.3 |
| Troponin I (ng/mL) | 0.00 | 0.01 | 0.02 | 0.06 | 0.19 | 0.85 | 16.86 |
| White Blood Cell Count (x10E3/uL) | 2.4 | 4.5 | 6.4 | 9.2 | 13.1 | 17.8 | 32.5 |
| Diastolic blood pressure (mmHg) | 49.7 | 58.5 | 64.0 | 70.3 | 76.7 | 82.6 | 94.3 |
| SPO2 (%) | 87.1 | 91.8 | 93.4 | 95.1 | 96.9 | 98.3 | 99.9 |
| Systolic blood pressure (mmHg) | 93.3 | 105.5 | 114.0 | 125.6 | 139.2 | 151.9 | 173.0 |

